# MENTOR: An Evidence-Informed Large Language Model-Based Multi-Agent System for Efficient Selection of Institution-Specific Negative Control Outcomes

**DOI:** 10.64898/2026.11.10.26367768

**Authors:** Taeseok Yang, Junhyuk Chang, Yeonghyeon Kim, Jaehyeong Cho, Kyungseon Choi, Minkook Son, Chungsoo Kim

**Author notes:** Corresponding author: Chungsoo Kim, Pharm.D. Ph.D. Department of Biomedical Informatics, Ajou University School of Medicine, 164 Worldcup-ro, Hongjae Bd 507-1, Suwon, 16499, Republic of Korea.

## Abstract

**Objective:** Negative control outcomes (NCOs) help detect and correct systematic error in observational studies, yet their selection remains largely manual. The Common Evidence Model (CEM) can generate candidate lists, but researchers must still screen them for plausibility as true nulls and for sufficient observation in each institution’s data. We developed and evaluated MENTOR (Multi-agent Evidence-informed Negative conTrol Outcome selection with Reasoning), a multi-agent system of large language models (LLMs) that combines CEM evidence with LLM reasoning to select institution-specific NCOs.

**Methods:** Four LLM agents operate on an institution’s Observational Medical Outcomes Partnership common data model. A Curator summarizes drug labels, a Suggester assesses locally frequent candidates with CEM evidence and a first-pass LLM judgment, an Adjudicator resolves their disagreements, and a Diagnostician relaxes the patient-count threshold until a target number of NCOs is met. Using Qwen3.6-27B, we evaluated 349 candidates for three drug pairs in a Korean tertiary hospital’s electronic health records (2.9 million patients) against a blinded LLM judge from a different model family, validated by three experts on 90 candidates. We compared accuracy, positive predictive value (PPV), and sensitivity with CEM-only and first-pass-LLM-only selection.

**Results:** MENTOR proposed 243 NCOs versus 121 by CEM-only, with higher accuracy (0.83 [0.79–0.87] vs. 0.53 [0.48–0.58]), sensitivity (0.89 vs. 0.41), and PPV (0.86 vs. 0.79). Judge–expert agreement was κ=0.63. Adjudication, needed for 44.1% of candidates, added little accuracy, and only with reasoning mode. Concordant acceptances, which bypass it, contained about half of the false positives. Only 7%–9% of CEM-proposed outcomes had at least 10,000 patients at the institution.

**Conclusion:** Combining LLM judgments with CEM evidence and local data yielded more NCOs, with higher PPV, than CEM alone. Restricting reasoning to discordant candidates kept computation time below 40% of that with reasoning in all agents. Every decision comes with its evidence and rationale for investigators’ final review.

## 1. Introduction

A negative control outcome (NCO) is an outcome believed not to be causally affected by the exposure of interest [1, 2]. Because an NCO shares the bias structure of the outcome of interest but carries no true effect, any systematic departure from the null in its estimate may indicate residual confounding, selection bias, or information bias [1, 2]. This property underlies empirical calibration, in which effect estimates for a large set of NCOs are used to derive an empirical null distribution and to calibrate the p-values and confidence intervals for the outcome of interest [3, 4]. Yet in a scoping review of pharmacoepidemiologic studies that used negative controls, 81% used them to detect bias but only 9% to correct bias and 4% to calibrate p-values [5]. Nearly half (46%) relied on a single negative control [5]. Calibration requires a large set of NCOs to capture a diverse mix of biases and to stabilize the null distribution, with 50 to 100 suggested as a rule of thumb [6]. NCOs must be selected not only accurately but also in sufficient numbers.

In practice, however, NCOs are still selected largely by hand. The main automated resource is the Common Evidence Model (CEM) [7], developed by the Observational Health Data Sciences and Informatics (OHDSI) community. It proposes candidates for which no evidence of association has been documented. However, absence of evidence does not establish a plausible null, and CEM does not account for the data available at a given institution. Large language models (LLMs) can read drug information and reason about causal plausibility, but applying them to NCO selection raises three design requirements. First, because LLMs are prone to hallucination, their judgments should complement, not replace, the evidence that CEM integrates from product labels, spontaneous reports, and literature. Second, extended deliberative reasoning is computationally costly [8], so deliberation should be applied selectively to the candidates that need it. Third, selection should reflect local data availability, so that the resulting set is both usable at the institution and large enough for calibration. Evaluating such a system requires a reference standard validated by human experts [9].

In this study, we developed MENTOR (Multi-agent Evidence-informed Negative conTrol Outcome selection with Reasoning), a multi-agent system that selects institution-specific NCOs on each institution’s Observational Medical Outcomes Partnership (OMOP) common data model (CDM) [10]. The contributions of this work are threefold. First, a hybrid adjudication mechanism combines CEM evidence with LLM plausibility reasoning. A reasoning-enabled agent resolves only the candidates on which the two disagree, confining costly deliberation to discordant cases. Second, a search loop accounts for local data availability by relaxing the patient-count threshold until the target number of NCOs is reached. Third, an evaluation uses an LLM judge from a different model family, validated against domain experts.

### Statement of Significance

|  |  |
|---|---|
| **Problem or Issue** | Selecting negative control outcomes (NCOs) is labor-intensive. Empirical calibration requires dozens of NCOs [6], yet extensively verifying every candidate is impractical, and the resulting selections vary across investigators and studies. |
| **What is Already Known** | The Common Evidence Model (CEM) automates the previous manual search for NCO candidates, but it relies on counting co-occurrence evidence and does not assess semantic plausibility. It also does not consider institution-specific data characteristics, so many proposed outcomes may be unusable at a given institution. |
| **What this Paper Adds** | MENTOR, a multi-agent system that combines CEM evidence collection with LLM-based semantic reasoning, achieving a higher positive predictive value than CEM while proposing more NCOs. Costly deliberative reasoning is reserved for candidates on which CEM and the first-pass judgment disagree, and an iterative loop searches for a target number of NCOs. |
| **Who would benefit from the new knowledge in this paper** | Investigators conducting observational studies who use NCOs to detect residual confounding and other biases and to correct for them through empirical calibration. |

## 2. Related Work

### 2.1 NCO selection in observational research

NCO selection has remained largely ad hoc. Candidates are usually proposed based on investigators’ domain knowledge or existing literature. A recent scoping review of 42 observational studies of glucagon-like peptide-1 receptor agonists found that the stated rationale for selection rested most often on clinical or subject-matter reasoning, and no study discussed the assumptions behind its negative controls [11]. This approach is labor-intensive. It also offers limited reproducibility across investigators and studies. More fundamentally, the defining assumption of an NCO is that no causal relationship exists between the exposure and the outcome. This assumption must be checked for every candidate, which is rarely feasible by hand across the full space of potential outcomes.

### 2.2 Knowledge-based approaches to NCO selection

To alleviate this burden, OHDSI developed CEM [7], a knowledge base that extends an earlier automated repository of drug–adverse reaction evidence [12]. This precursor was used to identify the 76 NCOs of a multinational antihypertensive study [13]. CEM integrates evidence from product labels, spontaneous reporting systems, and the published literature, and proposes exposure–outcome pairs lacking documented evidence of association as NCO candidates [7, 12]. Even so, the final list is still curated by hand. Earlier studies reviewed prevalence-ranked candidates manually [14] or had two physicians reduce 690 automatically filtered pairs to about 100 [15].

Two aspects of NCO selection lie outside what CEM was designed to do, and both are currently left to manual review. The first is plausibility. CEM screens by the presence or absence of documented evidence. This narrows the candidate space efficiently but does not by itself establish that a null relationship is plausible. Absence of evidence may reflect an unstudied pair, and a single piece of evidence excludes an outcome even when that evidence arises from confounding by indication or from a loosely related parent concept. Because CEM aggregates co-occurring index tags, such as Medical Subject Headings, rather than reading the source text, this judgment falls to the reviewer. The reviewer must also account for under- and over-reporting in the underlying sources [16, 17]. The second is data availability. CEM is a database-independent knowledge base, so it does not consider whether a candidate outcome is sufficiently observed at a given institution. OMOP databases differ in coding granularity and practice despite shared standardized vocabularies, and institutions differ in patient characteristics and size [18, 19]. An NCO can be analyzed only if enough patients in the study cohorts experience the outcome, so a proportion of proposed NCOs may be unusable in a particular database. Multi-institutional studies typically fix a single NCO set for comparability across sites, and candidates with too few events are dropped at each site. Smaller institutions are therefore left with fewer NCOs for calibration.

### 2.3 LLMs in drug safety and multi-agent systems

LLMs are increasingly applied in medicine [20]. They can read and interpret drug information [21], judge whether a causal relationship is plausible [22], and invoke external tools to retrieve structured evidence [23]. Recent examples include AgentMD [24] and agents that construct OMOP concept sets [25]. However, LLMs can generate plausible but unsupported content [26], and how they weigh supplied evidence depends on how it relates to their parametric knowledge [27]. This argues for pairing LLM reasoning with evidence-based screening rather than replacing it. Extended reasoning improves performance on difficult problems but spends excessive computation on simple ones [28].

Multi-agent LLM systems divide a task among specialized agents to make such selective allocation possible. However, they fail in characteristic ways, such as flaws in system design, misalignment between agents, and missing or incorrect verification [29]. Long inputs add a further risk, because LLMs use information in the middle of a long context less reliably than at its beginning or end [30]. Evaluating such systems at scale increasingly relies on LLM judges. They can agree with human preferences about as well as humans agree with each other [31] but tend to favor their own generations [32]. This motivates using a judge from a different model family and validating it against human experts.

## 3. Methods

### 3.1 Multi-agent system

#### 3.1.1 Overview and design principles

MENTOR generates institution-specific NCOs from a database in the OMOP CDM. Its inputs are the ingredient-level OMOP concept IDs of the target and comparator drugs. Its output is a set of concept IDs for valid NCOs with rationales, exported as an ATLAS concept set expression and a CohortGenerator negative control outcome table (Supplementary S6). Four agents (Curator, Suggester, Adjudicator, and Diagnostician) are orchestrated as a directed graph (**Figure 1**). Candidates are drawn from a Concept Pool built from the institution’s own patient counts. Each candidate is assessed by two independent lines of evidence, CEM evidence and LLM reasoning over drug information, and labeled concordant or discordant according to whether the two agree. Only discordant candidates are passed to the Adjudicator. The Diagnostician then lowers the patient-count threshold (the minimum number of patients with the outcome; min_patients in the code) stepwise until a target number of NCOs is reached.

**Figure 1.**
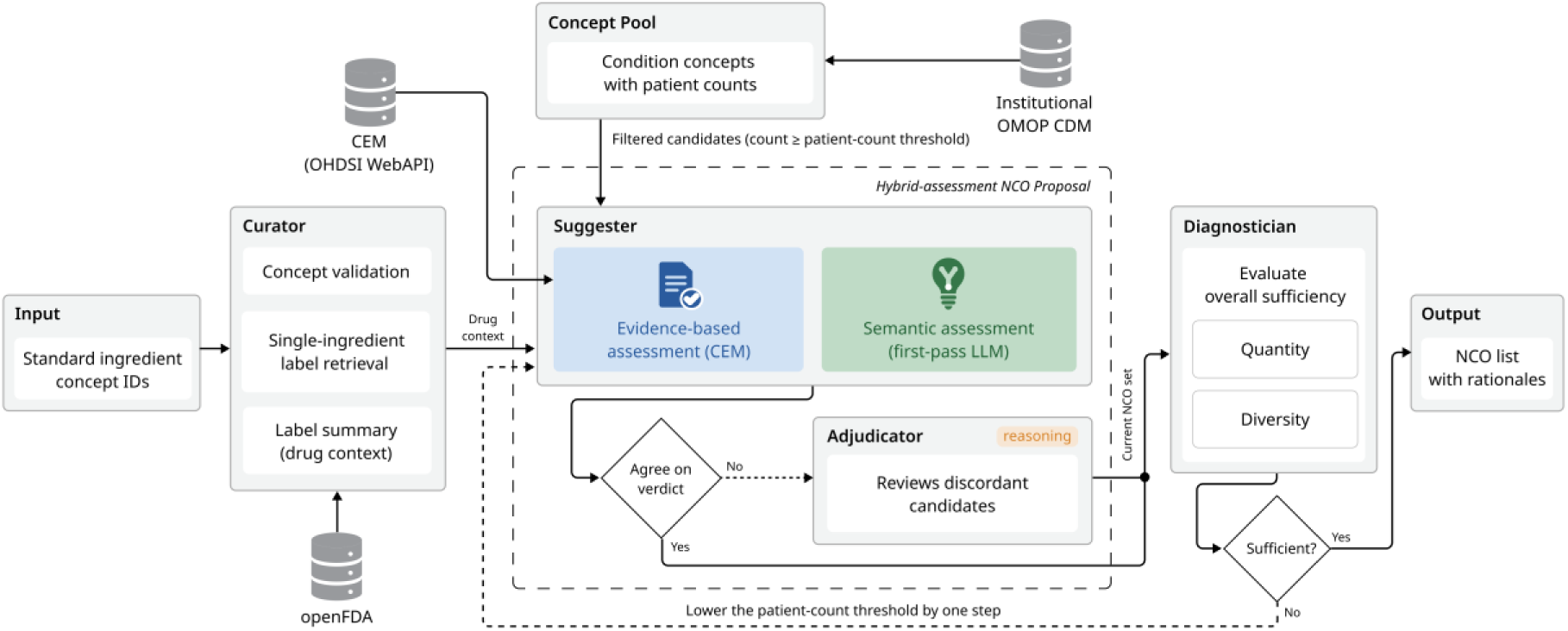
Architecture of MENTOR (Multi-agent Evidence-informed Negative conTrol Outcome selection with Reasoning). The diagram reads from left to right. Solid arrows show the main data flow, and dotted arrows show conditional paths. Each candidate from the Concept Pool is assessed with CEM evidence (blue) and by a first-pass LLM judgment (green). Candidates on which the two agree bypass the Adjudicator; only discordant candidates are reviewed by the Adjudicator, the only agent with reasoning mode enabled (orange label). The dashed box marks the steps repeated at each patient-count threshold. If the Diagnostician judges the NCO set insufficient, the threshold is lowered and the steps are repeated. CEM, Common Evidence Model; LLM, large language model; NCO, negative control outcome; OMOP CDM, Observational Medical Outcomes Partnership common data model.

#### 3.1.2 Curator

The Curator verifies that each input concept ID is a standard ingredient concept in the OMOP vocabulary and retrieves its name for label lookup. It then queries the openFDA drug labeling application programming interface (API) of the U.S. Food and Drug Administration [33] for single-ingredient labels whose substance or generic name matches the ingredient, falling back to all matching labels when no single-ingredient label exists. Because label text often differs across manufacturers, the Curator takes the text variant shared by the largest number of manufacturers. The sections used are indications, mechanism of action, contraindications, and four adverse-effect sections (**Supplementary S1**). An LLM, instructed not to add information absent from the labels, condenses this text into a structured drug context of standardized medical terms (**Supplementary S2**). This drug context informs all subsequent LLM judgments.

#### 3.1.3 Suggester

The Suggester first constructs the Concept Pool from the institution’s CDM by counting, for each standard condition concept, the distinct patients with at least one record of that concept or any of its descendants. Overly broad and pregnancy-related concepts are removed with a predefined exclusion list. The pool is built once per institution and reused across drug pairs.

Candidates meeting the current patient-count threshold are assessed in two ways. In the evidence-based assessment, the two ingredients are queried in CEM through the OHDSI WebAPI, and a candidate is proposed by CEM if no CEM source reports an association with either drug. Candidates not covered by CEM are outside the system’s scope. In the plausibility assessment (first pass), the LLM receives the drug context, the candidate’s name, and its ancestor concepts up to three levels. It returns a valid/invalid judgment with a two- to four-sentence rationale. All judging prompts share one NCO definition. An outcome is valid only if, for each drug, (1) it is not an indication, (2) it is not a contraindication or known adverse effect, and (3) it has no causal pathway through the drug’s mechanism, metabolism, or pharmacokinetics. The LLM may draw on its own pharmacological knowledge for criterion (3), but absence from the drug context alone is not treated as evidence of no relationship. A candidate is labeled concordant if the two assessments agree and discordant otherwise.

#### 3.1.4 Adjudicator

The Adjudicator resolves every discordant candidate with the model’s reasoning mode (thinking mode in Qwen3) enabled. It also resolves any candidate whose first-pass call returned no parsable judgment. Besides the first-pass inputs, it receives the CEM verdict and the first-pass judgment and rationale. It also receives the CEM evidence counts by source (publications, product labels, and spontaneous reports) and by matching level (exact, descendant, parent, and ancestor), and the CEM flags, such as indication or contraindication, drug-induced, or too broad (**Supplementary S3**). It is instructed to identify which source is likely wrong without default preference. Exact or descendant matches in product labels, and other exact or descendant evidence with a plausible pathway, are treated as disqualifying. Evidence found only at the parent or ancestor level, or attributable to confounding by indication, does not preclude acceptance. When uncertainty remains, the candidate is rejected, because an invalid NCO biases calibration more than an omitted one (**Supplementary S2.3**).

#### 3.1.5 Diagnostician

The Diagnostician checks whether the current NCO set has reached a target number (100 by default). Because a parent concept and its children, or near-synonymous concepts, may all be judged valid while largely capturing the same patients and bias, the raw number of NCOs overstates the number of independent NCOs. The Diagnostician therefore receives the names of the current NCOs and is instructed to estimate the effective number that would remain if each group of parent–child or near-synonymous concepts were counted as one. The LLM makes this estimate from the concept names rather than computing it from the vocabulary hierarchy. The effective number, not the raw number, is compared with the target. It is also instructed to judge diversity, that is, whether the set spans many organ systems, disease types, and clinical domains, and to treat a concentrated set as insufficient even if it is large. It returns a binary decision with a rationale (Supplementary S2.4). If the effective number falls short of the target, the threshold is lowered by one step, and the Suggester and Adjudicator process the newly eligible candidates. The loop ends when the target is reached or when the next threshold would fall below a predefined floor.

### 3.2 Experimental setup

#### 3.2.1 Data source and target drugs

We used the Ajou University School of Medicine (AUSOM) database, which contains the electronic health records of approximately 2.9 million patients with visits from January 1994 to November 2025, converted to the OMOP CDM (CDM version 5.3; OMOP standardized vocabularies v5.0, 27 February 2025). Three target–comparator pairs were selected from different therapeutic areas: (1) antidiabetic agents, empagliflozin (OMOP concept ID 45774751) versus sitagliptin (1580747); (2) anticoagulants, warfarin (1310149) versus apixaban (43013024); and (3) antihypertensive agents, lisinopril (1308216) versus hydrochlorothiazide (974166). Each pair shares an indication, so confounding by indication is a realistic source of spurious evidence.

#### 3.2.2 CEM baseline

We obtained CEM-based NCO candidates through the WebAPI of OHDSI ATLAS [34] (public instance; WebAPI 2.14.0; queried in October 2026; **Supplementary S4**). The WebAPI proposes an exposure–outcome pair only if no source reports an association. ATLAS also applies an optimization step that marks an outcome as not proposed when one of its ancestors is proposed. Because this step reflects redundancy rather than evidence, such outcomes were treated as proposed in both MENTOR and the baseline. CEM was chosen as the baseline because it is the established automated resource for NCO selection.

#### 3.2.3 Candidate pool construction

The exclusion list is fixed and shared across institutions. It contains 15,951 concepts that are unsuitable as NCOs regardless of the drug pair. These are overly broad or nonspecific concepts and pregnancy-related concepts, whose occurrence is governed by reproductive status rather than drug exposure (**Supplementary S5**).

In AUSOM, 19,231 standard condition concepts were recorded for at least one patient, and 13,009 remained after exclusion. To evaluate all methods on a common candidate set, the experiments used a single fixed patient-count threshold of 10,000 without the threshold-lowering loop, which yielded 186 condition concepts. After excluding those not covered by CEM, 117, 115, and 117 candidates remained for the antidiabetic, anticoagulant, and antihypertensive pairs, respectively (349 in total; **Supplementary S5**).

#### 3.2.4 Models and experimental instantiation

MENTOR was implemented with LangGraph [35] (Supplementary S6). All agents used Qwen3.6-27B with FP8 quantization, served by vLLM 0.23.0 [36] on two NVIDIA RTX A6000 GPUs with tensor parallelism, at a temperature of 0. In the main configuration, reasoning mode was enabled for the Adjudicator only. Each configuration was run once per drug pair with the threshold and the floor fixed at 10,000, so that all configurations shared the same candidates and the Diagnostician was invoked once per run. The threshold-lowering loop from 70,000 down to 10,000 was reconstructed offline from these runs. The hyperparameters and prompts are given in Supplementary S1 and S2. The prompts were developed iteratively by the authors and frozen before these experiments.

#### 3.2.5 Reference standard

The primary reference standard was an LLM judge, Claude Opus 5 (Anthropic; model identifier claude-opus-5), from a different model family from the MENTOR model to mitigate self-preference bias [31, 32]. The judge was blinded to the CEM verdict and all MENTOR outputs. It received the candidate outcome, the drug context, and the same NCO definition as MENTOR. It was instructed to check the three conditions for each drug and to draw on its own pharmacological knowledge, because the drug context may be incomplete (**Supplementary S2.5**). Each candidate was judged once.

As a secondary reference, 90 candidates (30 per drug pair) were drawn from the evaluation set by stratified random sampling with proportional allocation. Three epidemiologists with 7–10 years of experience each reviewed one drug pair. Reviewers were blinded to the CEM verdicts, MENTOR outputs, and LLM-judge labels and followed a written labeling guideline (**Supplementary S7**). They were given only the drug pair and the outcome name but could consult pharmacological references, product labels, and the literature. They classified each candidate as a valid NCO, not a valid NCO, or indeterminate, with a brief reason. Reviewers were instructed not to use generative AI for the labeling decisions, although its use for translating or summarizing literature was permitted.

#### 3.2.6 Performance metrics and statistics

Each candidate was treated as a binary classification instance with a valid NCO as the positive class. Because invalid NCOs contaminate the empirical null distribution, positive predictive value (PPV) is the key safety metric, whereas sensitivity determines how many usable NCOs are obtained. We report the number of proposed NCOs, accuracy, PPV, negative predictive value (NPV), sensitivity, specificity, and Cohen’s κ [37], with 95% confidence intervals (CIs) from percentile bootstrap (2,000 resamples, stratified by drug pair for pooled estimates). Paired differences in correctness were tested with the exact McNemar test.

Estimates are reported against the judge labels on the full evaluation set and against the human labels on the human-labeled sample, excluding indeterminate ratings. Consistency between the two references was assessed by the Spearman rank correlation of accuracies across all methods and configurations and by the direction of each configuration’s difference from the main configuration. The judge was validated against the human labels by percent agreement, Cohen’s κ, sensitivity, specificity, PPV, and NPV, with an exact McNemar test for asymmetric disagreement. In sensitivity analyses, indeterminate ratings were treated as valid or not valid.

#### 3.2.7 Secondary analyses

*(a) Component analysis*. We compared four decision rules for discordant candidates, namely CEM only, first-pass LLM only, the Adjudicator without reasoning mode, and the Adjudicator with reasoning mode (the main configuration). Configuration ablations changed one factor at a time. They enabled reasoning mode in all agents, removed the drug context (ingredient names only), used raw instead of summarized label text, or removed the concept hierarchy. Agreement between configurations was summarized by the Jaccard index of the final NCO sets. Reproducibility was assessed by comparing first-pass outputs between the two Adjudicator configurations, which share identical Curator and Suggester settings.
*(b) Discordance analysis, adjudication load, and cost*. We partitioned the MENTOR output into concordant and discordant candidates and counted false positives (FPs) in each group against the judge labels. For concordant candidates, which bypass adjudication, the error rate was also estimated from the human-labeled sample. Adjudication load was the number and proportion of candidates routed to the Adjudicator. LLM calls, tokens (input, output, and reasoning), and wall-clock time per agent were extracted from the run logs. Time spent waiting for the CEM WebAPI was recorded separately.
*(c) Availability and threshold sensitivity*. We counted the outcomes in the CEM candidate list obtained through the WebAPI that had at least one, 1,000, 5,000, and 10,000 patients in AUSOM (with descendants, without the exclusion list). Because candidates are judged independently of the threshold, the NCO sets the loop would produce at higher thresholds are subsets of the set at 10,000. We therefore reconstructed from the main runs the numbers of NCOs proposed by MENTOR and by CEM, and the MENTOR PPV, at thresholds from 70,000 to 10,000 in steps of 1,000.
*(d) Error analysis*. All misclassified candidates in the main configuration (FPs and false negatives [FNs] relative to the judge) were coded with a fixed codebook (Supplementary S8, S9). The codes covered decision path, error type (whether the evidence needed for the correct decision was in the drug context), and failure mode in the deciding agent’s rationale. Codes were assigned by Claude Opus 5.5 (claude-opus-5-5), a model different from both the MENTOR model and the judge, and were not verified by human reviewers.

#### 3.2.8 Reporting and ethics

This study is reported in accordance with the TRIPOD-LLM checklist [38] (Supplementary S10). The Institutional Review Board of Ajou University Hospital reviewed the study and granted an exemption (AJOUIRB-EX-2026-412), waiving the requirement for informed consent because only de-identified data were analyzed retrospectively. Patient-level data were accessed only within the institutional environment. The self-hosted LLM received only concept names, drug information, and aggregate patient counts, and external services (openFDA, the CEM WebAPI, and the LLM APIs used for the judge and error coding) received no patient data.

## 4. Results

### 4.1 Candidates and reference labels

All 349 candidates met the patient-count threshold of 10,000, with counts ranging from 10,144 to 187,182 (median [interquartile range], 17,262 [12,861–24,392]). The LLM judge labeled 233 (66.8%) as valid NCOs (69 [59.0%], 90 [78.3%], and 74 [63.2%] for the antidiabetic, anticoagulant, and antihypertensive pairs, respectively). Of the 90 sampled candidates, the human experts rated 49 (54.4%) as valid, 14 (15.6%) as not valid, and 27 (30.0%) as indeterminate (6, 13, and 8 in the pairs, respectively). This left 63 for the primary analyses, of which 49 (77.8%) were valid.

### 4.2 Validation of the LLM judge

On the 63 human-labeled candidates, the LLM judge achieved an accuracy of 0.86 [95% CI, 0.76–0.94], a sensitivity of 0.86 [0.75–0.95], a specificity of 0.86 [0.64–1.00], a PPV of 0.95 [0.89–1.00], an NPV of 0.63 [0.41–0.86], and a Cohen’s κ of 0.63 [0.39–0.83], corresponding to substantial agreement [39] (Supplementary S11). Of the nine disagreements, seven were cases in which the LLM judge rejected outcomes that the expert accepted, and two were vice versa (McNemar P=0.18). In the primary analysis, agreement was highest for empagliflozin–sitagliptin (κ=0.75) and lower for warfarin–apixaban (κ=0.45) and lisinopril–hydrochlorothiazide (κ=0.42). Four of the seven conservative disagreements were in the lisinopril–hydrochlorothiazide pair, two of which cited mechanistic pathways such as thiazide effects on calcium balance (Supplementary S12). In the sensitivity analyses, treating indeterminate ratings as valid or as not valid gave κ values of 0.46 and 0.38 and PPVs of 0.97 and 0.68, respectively.

### 4.3 NCO selection performance

MENTOR proposed 243 NCOs (69.6% of candidates), compared with 121 (34.7%) proposed by CEM (Figure 2a, Table 1; candidate-level results in Supplementary S13). Pooled accuracy was 0.83 [95% CI, 0.79–0.87] for MENTOR and 0.53 [0.48–0.58] for CEM (McNemar P<0.001). Sensitivity was 0.89 [0.85–0.93] vs. 0.41 [0.35–0.47] and NPV 0.76 [0.68–0.84] vs. 0.39 [0.33–0.46]. PPV was also higher for MENTOR (0.86 [0.81–0.90] vs. 0.79 [0.71–0.86]; difference, 0.071 [0.019 to 0.127]), whereas specificity was lower (0.70 [0.62–0.78] vs. 0.78 [0.70–0.85]). The accuracy advantage of MENTOR was observed in every drug pair and was largest for warfarin–apixaban. In this pair, CEM proposed 30 outcomes (sensitivity, 0.31) and MENTOR proposed 87 (sensitivity, 0.92), with PPVs of 0.93 and 0.95, respectively.

**Figure 2.**
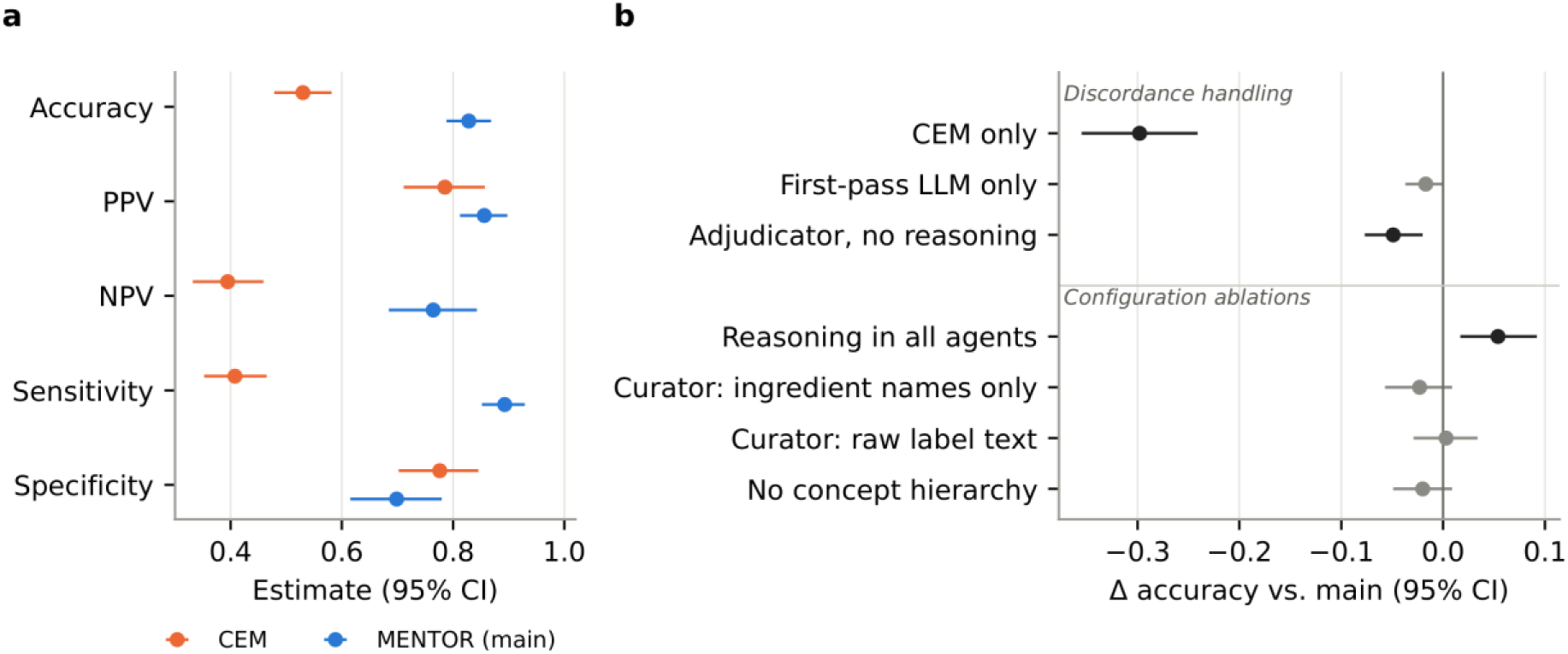
(a) Pooled performance of MENTOR (main configuration, with reasoning mode in the Adjudicator only) and CEM against the LLM judge (n = 349). (b) Difference in accuracy between each alternative and the main configuration (alternative minus main); negative values indicate lower accuracy than the main configuration. Discordance handling replaces the Adjudicator’s decision on discordant candidates, and configuration ablations change one setting at a time. Black markers indicate 95% CIs that exclude zero, and gray markers indicate CIs that include zero. Error bars are bootstrap 95% CIs stratified by drug pair. CEM, Common Evidence Model; CI, confidence interval; LLM, large language model; NPV, negative predictive value; PPV, positive predictive value.

**Table 1.**
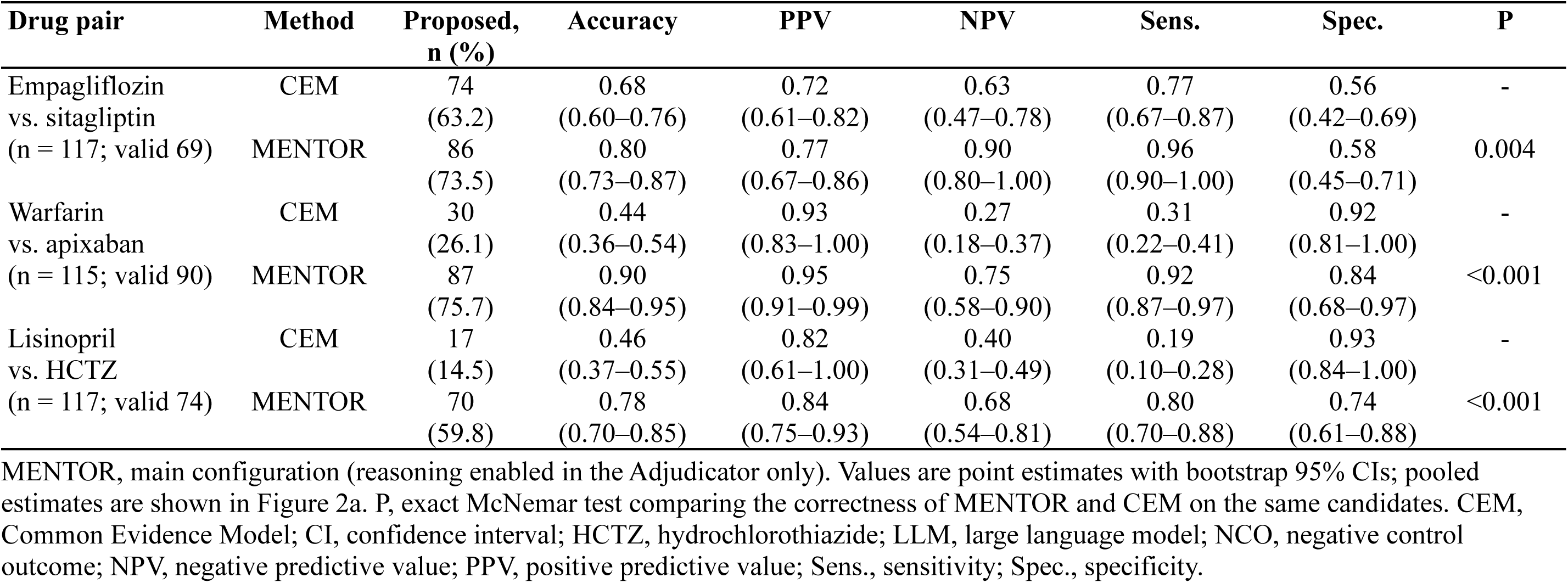
NCO selection performance of MENTOR and CEM by drug pair against the LLM judge.

| Drug pair | Method | Proposed, n (%) | Accuracy | PPV | NPV | Sens. | Spec. | P |
|---|---|---|---|---|---|---|---|---|
| Empagliflozin<br>vs. sitagliptin<br>(n = 117; valid 69) | CEM | 74 (63.2) | 0.68 (0.60–0.76) | 0.72 (0.61–0.82) | 0.63 (0.47–0.78) | 0.77 (0.67–0.87) | 0.56 (0.42–0.69) | - |
|  | MENTOR | 86 (73.5) | 0.80 (0.73–0.87) | 0.77 (0.67–0.86) | 0.90 (0.80–1.00) | 0.96 (0.90–1.00) | 0.58 (0.45–0.71) | 0.004 |
| Warfarin<br>vs. apixaban<br>(n = 115; valid 90) | CEM | 30 (26.1) | 0.44 (0.36–0.54) | 0.93 (0.83–1.00) | 0.27 (0.18–0.37) | 0.31 (0.22–0.41) | 0.92 (0.81–1.00) | - |
|  | MENTOR | 87 (75.7) | 0.90 (0.84–0.95) | 0.95 (0.91–0.99) | 0.75 (0.58–0.90) | 0.92 (0.87–0.97) | 0.84 (0.68–0.97) | <0.001 |
| Lisinopril<br>vs. HCTZ<br>(n = 117; valid 74) | CEM | 17 (14.5) | 0.46 (0.37–0.55) | 0.82 (0.61–1.00) | 0.40 (0.31–0.49) | 0.19 (0.10–0.28) | 0.93 (0.84–1.00) | - |
|  | MENTOR | 70 (59.8) | 0.78 (0.70–0.85) | 0.84 (0.75–0.93) | 0.68 (0.54–0.81) | 0.80 (0.70–0.88) | 0.74 (0.61–0.88) | <0.001 |

MENTOR, main configuration (reasoning enabled in the Adjudicator only). Values are point estimates with bootstrap 95% CIs; pooled estimates are shown in Figure 2a. P, exact McNemar test comparing the correctness of MENTOR and CEM on the same candidates. CEM, Common Evidence Model; CI, confidence interval; HCTZ, hydrochlorothiazide; LLM, large language model; NCO, negative control outcome; NPV, negative predictive value; PPV, positive predictive value; Sens., sensitivity; Spec., specificity.

On the 63 human-labeled candidates, MENTOR outperformed CEM (accuracy, 0.79 vs. 0.44; PPV, 0.91 vs. 0.79). Across the eight systems compared (CEM, the first pass, and six MENTOR configurations), accuracy rankings under human labels correlated with rankings under LLM judge labels, both on the same 63 candidates (Spearman ρ=0.98, P<0.001) and on the full set of 349 (ρ=0.84, P=0.009).

### 4.4 Component analysis

Two runs with identical Curator and Suggester settings produced identical drug contexts and first-pass judgments, including rationales, for all 349 candidates (Supplementary S14). Among the discordance-handling strategies, the Adjudicator with reasoning mode achieved the highest accuracy (Figure 2b; Supplementary S15), although its gain over the first pass alone was not statistically significant (0.017 [0.000 to 0.037], P=0.11). The gain came from sensitivity (0.021 [0.004 to 0.042]) and NPV (0.037 [0.009 to 0.072]), with no difference in PPV (0.007 [−0.009 to 0.022]) or specificity (0.009 [−0.028 to 0.045]; Supplementary S15). Without reasoning mode, the Adjudicator rejected more candidates and performed worse than the first pass (−0.032 [−0.060 to −0.003], P=0.04). Reasoning mode therefore added 0.049 [0.020 to 0.077] (P=0.002). Among the 154 discordant candidates, accuracy was 0.18 when following CEM, 0.82 when following the first pass, 0.86 for the Adjudicator with reasoning mode, and 0.75 for the Adjudicator without reasoning mode (Supplementary S15).

In the ablation analysis, reasoning mode in all agents increased accuracy (0.054 [0.017 to 0.092], P=0.008) at 2.6 times the wall-clock time. Ingredient names only (−0.023 [−0.057 to 0.009], P=0.23), raw label text (P>0.99; 3.4-fold token usage), and no concept hierarchy (P=0.23) had no significant effect. Against the expert labels, the configurations ranked in the same direction relative to the main configuration, except that removing the concept hierarchy was slightly better (by two candidates) and using ingredient names only was tied. The final NCO sets overlapped substantially across configurations (Jaccard index, 0.83–0.90; Supplementary S16).

### 4.5 Concordance, false positives, and adjudication load

CEM and the first-pass judgment agreed on 195 candidates (55.9%) and disagreed on 154 (44.1%). The proportion of discordant candidates was 22.2%, 59.1%, and 51.3% for the antidiabetic, anticoagulant, and antihypertensive pairs, respectively (Supplementary S17, S18). Discordance was dominated by candidates that CEM rejected but the first pass accepted (136 of 154), of which the Adjudicator accepted 133 (97.8%). Of the 19 such candidates that the judge rated not valid, the Adjudicator rejected only 3. Of the 18 candidates that CEM accepted but the first pass rejected, the Adjudicator accepted 7 (38.9%).

Of the 35 FPs, 17 (48.6%) were concordant acceptances, which bypass adjudication. Of the 26 outcomes CEM proposed that the judge rated not valid, the first pass rejected only 9. The PPV was 0.83 (86/103) for concordant and 0.87 (122/140) for adjudicated acceptances. Of the 92 concordant rejections, 21 were valid according to the judge. Among the 63 expert-labeled candidates, 4 of 21 concordant acceptances and none of 23 adjudicated acceptances were rated not valid.

The main configuration used 512 LLM calls and 1.37 million tokens and took 158 minutes for the three pairs (53 minutes per pair, of which 2.6 minutes in total were spent waiting for CEM). The Adjudicator accounted for 30.1% of calls, 52.0% of tokens, and 82.3% of wall-clock time, largely because of reasoning tokens. Per proposed NCO, the system used 5,626 tokens and 38 seconds (Supplementary S19, S20).

### 4.6 Availability of usable NCOs

CEM, queried through ATLAS, proposed 227, 169, and 170 outcomes for the antidiabetic, anticoagulant, and antihypertensive pairs. Of these, 149 (65.6%), 106 (62.7%), and 100 (58.8%) had at least one patient in AUSOM, and only 20 (8.8%), 14 (8.3%), and 12 (7.1%) had at least 10,000 (Figure 3a; Supplementary S21). These are the raw WebAPI counts, before restoring optimization-removed descendants and applying the exclusion list, and thus differ from the 121 proposals evaluated above. As the threshold was lowered from 70,000 to 10,000, MENTOR reached 50 proposed NCOs (before collapsing related concepts) at 16,000, 16,000, and 14,000 patients, whereas CEM reached 50 only for empagliflozin–sitagliptin and at most 30 and 17 for the other pairs (Figure 3b). The MENTOR PPV was 0.69, 1.00, and 0.88 at 20,000 patients and 0.77, 0.95, and 0.84 at 10,000 (Supplementary S21). In all 18 runs, the Diagnostician judged the set at 10,000 (55–100 NCOs; estimated effective counts, approximately 40–60) insufficient for the target of 100.

**Figure 3.**
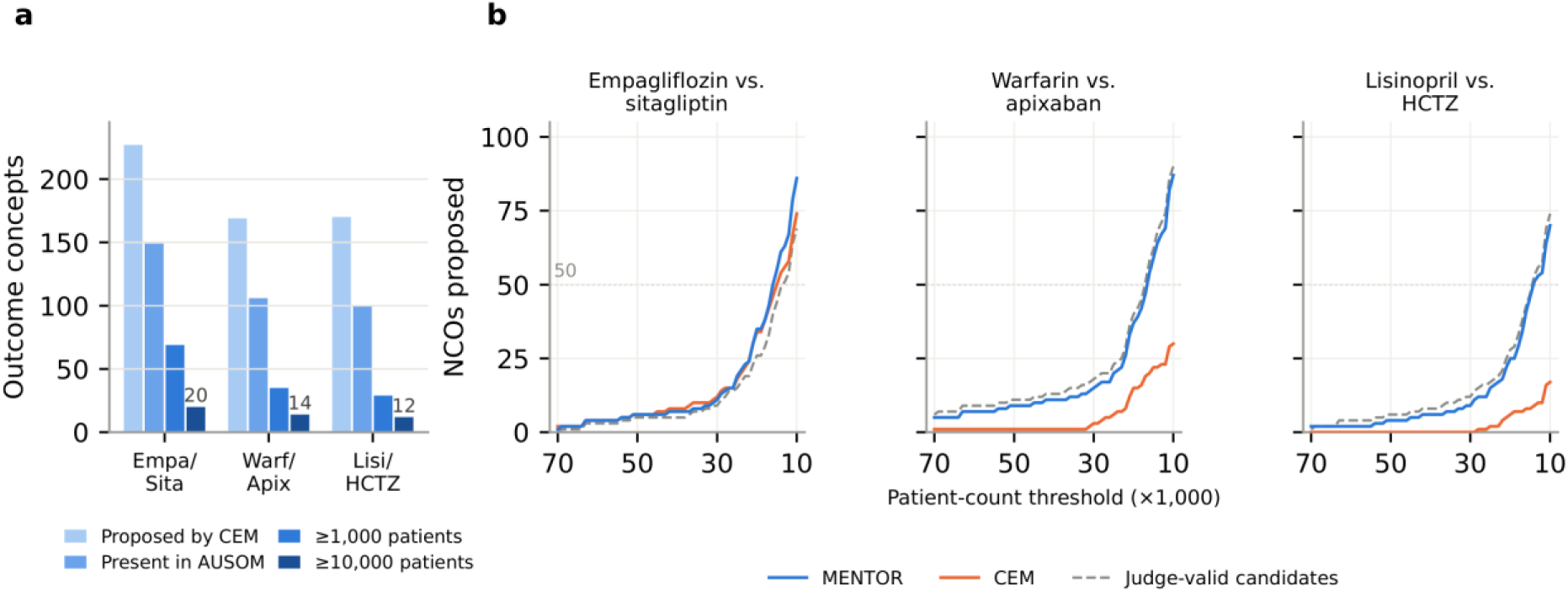
(a) Outcomes in the final CEM proposal list for each drug pair (after ATLAS optimization) and the numbers present in the AUSOM database (any patient, counted with descendants) and with at least 1,000 or 10,000 patients. (b) Number of NCOs proposed by MENTOR and by CEM (Section 3.2.2 definition), and number of judge-valid candidates, among the evaluated candidates as the patient-count threshold is lowered from 70,000 to 10,000 (reconstructed from the main runs). The dotted line marks 50 NCOs, the lower end of the 50–100 suggested in [6]. Empa/Sita, empagliflozin vs. sitagliptin; Warf/Apix, warfarin vs. apixaban; Lisi/HCTZ, lisinopril vs. hydrochlorothiazide; AUSOM, Ajou University School of Medicine; CEM, Common Evidence Model; HCTZ, hydrochlorothiazide; NCO, negative control outcome.

### 4.7 Error analysis

The main configuration produced 35 FPs and 25 FNs (Supplementary S18, S22). FPs arose about equally from adjudication and concordant acceptance (n=18 vs. 17). Most were label-grounded, that is, inferable from the drug context (n=24, 13 through two or more inference steps). None involved an outcome named verbatim in the context. The remaining 11 rested on established (n=5) or speculative (n=6) evidence outside it (Supplementary S9).

Among the 18 adjudicated FPs, the most frequent failure mode was mechanism-first filtering (n=7), in which an outcome was accepted because no direct mechanistic pathway was found, although a listed adverse effect could cause it indirectly. All seven involved rib fracture concepts, across all three drug pairs. Among 17 concordant FPs, 4 reflected taxonomic mismatch, in which a broader listed term (e.g., upper respiratory tract infection for acute sinusitis) was not linked to the outcome despite appearing in the hierarchy input. Three reflected label dismissal, in which a listed adverse effect was acknowledged but judged non-causal. Most FNs (18 of 25) reflected over-extended mechanistic reasoning, such as rejecting gastritis for warfarin–apixaban because of bleeding risk.

## 5. Discussion

In this study, a multi-agent LLM system informed by CEM evidence proposed about twice as many NCOs as CEM from the same institution-specific candidates, with higher accuracy, sensitivity, and PPV. The conclusion held under expert labels. Deliberative adjudication restricted to discordant candidates added a modest gain over the first-pass judgment, but only when the adjudicating model was allowed to reason. Finally, at the 10,000-patient threshold used here, only 7%–9% of CEM proposals were usable in the institution’s data.

CEM’s conservative yield follows from its screening rule. It rejects an outcome whenever any source reports evidence at any matching level, without distinguishing a causal effect. Most discordance (136 of 154) arose from such exclusions, and the Adjudicator accepted 133 of them, in agreement with the first pass. Concordance, however, did not guarantee correctness. The first pass identified only 9 of CEM’s 26 invalid proposals, and concordant acceptances accounted for about half of all FPs. First, selective deliberation concentrates reasoning where the two sources conflict and reduces computation relative to reasoning in all agents. It improves accuracy little, because the Adjudicator rarely overrode the first pass. Its contribution lies instead in producing rationales that cite the source and level of evidence, which a reviewer can audit. Second, enabling reasoning in all agents improved accuracy, but concordant acceptances still bypassed adjudication. Directing additional review to them is one way to improve the system.

The system is intended to support, not replace, the final review by investigators. The system automates much of this curation and returns, for every candidate, the CEM evidence by source and matching level and the first-pass rationale. For discordant candidates, it also returns the Adjudicator’s decision and reasoning. Investigators can therefore confirm the final list by checking the stated evidence and reasoning rather than reviewing every candidate from scratch. This review should include concordant acceptances, which contained about half of the FPs.

The availability analysis supports institution-specific selection. Of CEM’s proposals, 34%–41% did not occur in our database and only 7%–9% reached 10,000 patients. These counts precede the washout or time-at-risk restrictions that further reduce the events available in an actual study cohort. Because MENTOR operates on each institution’s own Concept Pool, it could run locally at each site of a distributed network and share only the resulting concept sets. Such local selection could complement a common network set and help restore sufficient numbers at sites where the common set falls short. At the 10,000-patient floor, no configuration in our data reached 100 effective NCOs, the upper end of the 50–100 suggested for calibration [6]. Whether that target is attainable at lower thresholds, and at what cost in PPV, remains to be examined.

Apart from the final investigator review, the system requires no manual steps. From ingredient concept IDs, it produces outputs that can be loaded directly into ATLAS or Strategus pipelines. A run took about 53 minutes per drug pair on a single self-hosted model, or 12 minutes with the Adjudicator’s reasoning mode disabled, but with lower sensitivity (0.80 vs. 0.89; Supplementary S15).

Most remaining errors were failures of reasoning over information the system already had, and four patterns suggest remedies. First, the Adjudicator ruled out downstream consequences, such as fractures from listed causes of falls, on mechanistic grounds without consulting the listed adverse effects. An explicit check of listed effects before mechanistic reasoning would address this. Second, some FPs dismissed an acknowledged adverse effect as non-causal, contrary to the NCO definition, which an explicit prompt rule could prevent. Third, concordant FPs missed broader listed terms present in the hierarchy input. Although limited to three levels, that input listed a median of 24 (up to 69) terms per candidate, including synonyms, grouping terms from other vocabularies, and very broad concepts such as ‘Infectious disease’. This may also explain why removing it had no significant effect. Fourth, most FNs rejected a valid outcome through a long or speculative causal chain. Bounding the length of admissible chains is one possible remedy. These remedies were not evaluated, because revising prompts on the same drug pairs would overfit.

That neither removing nor expanding the drug labels changed accuracy suggests that the model relied largely on its own pharmacological knowledge. This is consistent with reports that LLMs adopt supplied evidence depending on how it relates to their parametric knowledge [27] and use long inputs unevenly [30]. Whether the drug context matters more for less familiar drugs than the six studied here remains open.

The evaluation relied on an LLM judge, which raises the question of circularity. To mitigate it, we used a model from a different family, blinded it to the system outputs, and validated it against domain experts. Agreement was substantial (κ=0.63), and the judge’s PPV was high (0.95). Its disagreements tended to be conservative, which would bias the system’s PPV downward, and the main conclusions held on expert labels.

This study has further limitations. In terms of generalizability, it was conducted in a single institution with three drug pairs. Applicability to other institutions and drug classes remains to be shown. The drug context depends on U.S. product labels, so drugs or indications not approved in the United States are not covered. For experimental design, we restricted candidates to concepts with at least 10,000 patients and an existing record in CEM so that all methods could be compared on the same candidates. We also reconstructed the threshold-lowering loop offline instead of running it. Performance on rarer outcomes and on outcomes outside CEM coverage, and the behavior of the loop below 10,000 patients, are unknown. Each configuration was run once with a single LLM. The architecture does not depend on a specific model, but other models were not evaluated. Because the prompts were developed on the same drug pairs, the results may favor the system. In terms of the reference standard, the system and the judge share the NCO definition and the drug context. The judge validates adherence to that definition rather than an independent conception of validity. Agreement was also weakest for lisinopril–hydrochlorothiazide (κ=0.42). The human sample was small, contained 30.0% indeterminate ratings, and had one reviewer per pair, so inter-rater agreement could not be assessed. Reviewers were instructed not to use generative AI, but individual use could not be fully controlled. How much the system shortens investigator review remains to be evaluated. The error analysis was coded by a third model with a fixed codebook but was not verified by human reviewers. Finally, the Diagnostician only judges whether the current set is sufficient. It does not remove redundant concepts or optimize the final set for diversity, which is left to investigators.

## 6. Conclusion

We developed a multi-agent LLM system that selects institution-specific negative control outcomes. It combines CEM evidence with LLM reasoning over drug information and accounts for data availability in each institution’s OMOP CDM. In a single-institution evaluation of three drug pairs, the system proposed substantially more NCOs than CEM with higher sensitivity and PPV. The system is intended as a decision-support tool whose final list investigators confirm against the evidence and rationales it provides. Adjudication of discordant candidates helped only when reasoning mode was enabled. The system requires only standard OHDSI resources and a locally served open-weight LLM. It could therefore be run at other institutions with an OMOP CDM, including each site of a network study, without patient-level data leaving the site. Future work should evaluate the system across multiple institutions and in real-world study workflows.

## Supporting information

Supplementary Meterials

Error coding sheet

## Data Availability

Patient-level data from the Ajou University School of Medicine (AUSOM) database cannot be shared because of institutional data protection policies. Candidate-level outputs of all configurations, the judge labels, and the expert labels are provided in the Supplementary Data. The code is available at https://github.com/ABMI/MENTOR.

## Author Contributions

**Taeseok Yang:** Conceptualization, Methodology, Software, Formal analysis, Investigation, Data curation, Visualization, Writing – original draft; **Junhyuk Chang:** Writing – review & editing; **Yeonghyeon Kim:** Writing – review & editing; **Jaehyeong Cho:** Validation, Writing – review & editing; **Kyungseon Choi:** Validation, Writing – review & editing; **Minkook Son:** Validation, Writing – review & editing; **Chungsoo Kim:** Conceptualization, Methodology, Supervision, Project administration, Funding acquisition, Writing – review & editing.

## Data Availability

Patient-level data from the AUSOM database cannot be shared because of institutional data protection policies. Candidate-level outputs of all configurations, the judge labels, and the expert labels are provided in the Supplementary Data.

## Code Availability

The code for this study is available on GitHub at https://github.com/ABMI/MENTOR.

## Declaration of generative AI and AI-assisted technologies in the manuscript preparation process

During the preparation of this work, the authors used Claude (Anthropic) to draft, edit, and improve language and readability of portions of the manuscript. After using this tool, the authors reviewed and revised the content as necessary. The authors have full responsibility for the content of this manuscript.

## References

[1] M. Lipsitch, E. Tchetgen Tchetgen, T. Cohen, Negative controls: a tool for detecting confounding and bias in observational studies, Epidemiology 21 (2010) 383–388. 10.1097/EDE.0b013e3181d61eeb.

[2] Q. Yang, Z. Yang, X. Cai, H. Zhao, J. Jia, F. Sun, Advances in methodologies of negative controls: a scoping review, J. Clin. Epidemiol. 166 (2024) 111228. 10.1016/j.jclinepi.2023.111228.

[3] M.J. Schuemie, P.B. Ryan, W. DuMouchel, M.A. Suchard, D. Madigan, Interpreting observational studies: why empirical calibration is needed to correct p-values, Stat. Med. 33 (2014) 209–218. 10.1002/sim.5925.

[4] M.J. Schuemie, G. Hripcsak, P.B. Ryan, D. Madigan, M.A. Suchard, Empirical confidence interval calibration for population-level effect estimation studies in observational healthcare data, Proc. Natl. Acad. Sci. U.S.A. 115 (2018) 2571–2577. 10.1073/pnas.1708282114.

[5] Z. Zafari, J.-E. Park, C.H. Shah, S. dosReis, E.F. Gorman, W. Hua, Y. Ma, F. Tian, The state of use and utility of negative controls in pharmacoepidemiologic studies, Am. J. Epidemiol. 193 (2024) 426–453. 10.1093/aje/kwad201.

[6] M.J. Schuemie, Method validity, in: The Book of OHDSI, Observational Health Data Sciences and Informatics, 2021. https://ohdsi.github.io/TheBookOfOhdsi/MethodValidity.html (accessed 29 October 2026).

[7] Observational Health Data Sciences and Informatics, Common Evidence Model (CEM). https://github.com/OHDSI/CommonEvidenceModel/wiki (accessed 2 November 2026).

[8] S. Zhou, R. Ling, J. Chen, X. Wang, T. Fan, H. Wang, When more thinking hurts: overthinking in LLM test-time compute scaling, arXiv (2026). 10.48550/arXiv.2604.10739.

[9] L. Li, D. Li, C. Chen, R. Ma, R. Yu, M. Lin, R. Yin, L. Fan, C. Shyr, S. Ma, M. Liu, S. Bethard, LLM-as-a-judge in healthcare: a scoping analysis of applications, methods, and human alignment, arXiv (2026). 10.48550/arXiv.2605.25273.

[10] J.M. Overhage, P.B. Ryan, C.G. Reich, A.G. Hartzema, P.E. Stang, Validation of a common data model for active safety surveillance research, J. Am. Med. Inform. Assoc. 19 (2012) 54–60. 10.1136/amiajnl-2011-000376.

[11] Z. Li, Y. Huang, H. Chen, J. Li, How negative controls are used in pharmacoepidemiology: a methodological scoping review of real-world observational studies of glucagon-like peptide-1 receptor agonists, Pharmacoepidemiol. Drug Saf. 35 (2026) e70420. 10.1002/pds.70420.

[12] E.A. Voss, R.D. Boyce, P.B. Ryan, J. van der Lei, P.R. Rijnbeek, M.J. Schuemie, Accuracy of an automated knowledge base for identifying drug adverse reactions, J. Biomed. Inform. 66 (2017) 72–81. 10.1016/j.jbi.2016.12.005.

[13] M.A. Suchard, M.J. Schuemie, H.M. Krumholz, S.C. You, R. Chen, N. Pratt, C.G. Reich, J. Duke, D. Madigan, G. Hripcsak, P.B. Ryan, Comprehensive comparative effectiveness and safety of first-line antihypertensive drug classes: a systematic, multinational, large-scale analysis, Lancet 394 (2019) 1816–1826. 10.1016/S0140-6736(19)32317-7.

[14] M.J. Schuemie, P.B. Ryan, G. Hripcsak, D. Madigan, M.A. Suchard, Improving reproducibility by using high-throughput observational studies with empirical calibration, Philos. Trans. R. Soc. A Math. Phys. Eng. Sci. 376 (2018) 20170356. 10.1098/rsta.2017.0356.

[15] E.A. Voss, M.J. Schuemie, J. van der Lei, P.R. Rijnbeek, Assessing negative control exposure-outcome pair selection strategies on a replication study, Poster presented at: OHDSI Symposium, 2019. https://www.ohdsi.org/wp-content/uploads/2019/09/ERICA-VOSS_Assessing-Negative-Control-Exposure-Outcome-Pair-Selection-Strategies-on-a-Replication-Study_2019Symposium.pdf (accessed 29 October 2026).

[16] L. Hazell, S.A.W. Shakir, Under-reporting of adverse drug reactions: a systematic review, Drug Saf. 29 (2006) 385–396. 10.2165/00002018-200629050-00003.

[17] P.M. Cutroneo, D. Sartori, M. Tuccori, S. Crisafulli, V. Battini, C. Carnovale, C. Rafaniello, A. Capuano, E. Poluzzi, U. Moretti, E. Raschi, Conducting and interpreting disproportionality analyses derived from spontaneous reporting systems, Front. Drug Saf. Regul. 3 (2024) 1323057. 10.3389/fdsfr.2023.1323057.

[18] A. Ostropolets, C. Reich, P. Ryan, C. Weng, A. Molinaro, F. DeFalco, J. Jonnagaddala, S.-T. Liaw, H. Jeon, R.W. Park, M.E. Spotnitz, K. Natarajan, G. Argyriou, K. Kostka, R. Miller, A. Williams, E. Minty, J. Posada, G. Hripcsak, Characterizing database granularity using SNOMED-CT hierarchy, AMIA Annu. Symp. Proc. 2020 (2020) 983–992.

[19] E.A. Voss, R. Makadia, A. Matcho, Q. Ma, C. Knoll, M. Schuemie, F.J. DeFalco, A. Londhe, V. Zhu, P.B. Ryan, Feasibility and utility of applications of the common data model to multiple, disparate observational health databases, J. Am. Med. Inform. Assoc. 22 (2015) 553–564. 10.1093/jamia/ocu023.

[20] A.J. Thirunavukarasu, D.S.J. Ting, K. Elangovan, L. Gutierrez, T.F. Tan, D.S.W. Ting, Large language models in medicine, Nat. Med. 29 (2023) 1930–1940. 10.1038/s41591-023-02448-8.

[21] L. Wu, H. Fang, Y. Qu, J. Xu, W. Tong, Leveraging FDA labeling documents and large language model to enhance annotation, profiling, and classification of drug adverse events with AskFDALabel, Drug Saf. 48 (2025) 655–665. 10.1007/s40264-025-01520-1.

[22] E. Kıcıman, R. Ness, A. Sharma, C. Tan, Causal reasoning and large language models: opening a new frontier for causality, Trans. Mach. Learn. Res. (2024). 10.48550/arXiv.2305.00050.

[23] S. Yao, J. Zhao, D. Yu, N. Du, I. Shafran, K. Narasimhan, Y. Cao, ReAct: synergizing reasoning and acting in language models, in: Proc. 11th Int. Conf. Learn. Represent. (ICLR), 2023. 10.48550/arXiv.2210.03629.

[24] Q. Jin, Z. Wang, Y. Yang, Q. Zhu, D. Wright, T. Huang, N. Khandekar, N. Wan, X. Ai, W.J. Wilbur, Z. He, R.A. Taylor, Q. Chen, Z. Lu, AgentMD: empowering language agents for risk prediction with large-scale clinical tool learning, Nat. Commun. 16 (2025) 9377. 10.1038/s41467-025-64430-x.

[25] H. Chen, X. He, H. Dai, Y. Huang, M. Liu, J. Bian, Agentic authoring of OMOP concept sets from natural language, medRxiv (2026). 10.64898/2026.06.02.26354704.

[26] Z. Ji, N. Lee, R. Frieske, T. Yu, D. Su, Y. Xu, E. Ishii, Y.J. Bang, A. Madotto, P. Fung, Survey of hallucination in natural language generation, ACM Comput. Surv. 55 (2023) 248. 10.1145/3571730.

[27] J. Xie, K. Zhang, J. Chen, R. Lou, Y. Su, Adaptive chameleon or stubborn sloth: revealing the behavior of large language models in knowledge conflicts, in: Proc. 12th Int. Conf. Learn. Represent. (ICLR), 2024. 10.48550/arXiv.2305.13300.

[28] X. Chen, J. Xu, T. Liang, Z. He, J. Pang, D. Yu, L. Song, Q. Liu, M. Zhou, Z. Zhang, R. Wang, Z. Tu, H. Mi, D. Yu, Do NOT think that much for 2+3=? On the overthinking of long reasoning models, in: Proc. 42nd Int. Conf. Mach. Learn. (ICML), PMLR 267, 2025: pp. 9487–9499. 10.48550/arXiv.2412.21187.

[29] M. Cemri, M.Z. Pan, S. Yang, L.A. Agrawal, B. Chopra, R. Tiwari, K. Keutzer, A. Parameswaran, D. Klein, K. Ramchandran, M. Zaharia, J.E. Gonzalez, I. Stoica, Why do multi-agent LLM systems fail?, in: Adv. Neural Inf. Process. Syst. 38 (NeurIPS 2025), Datasets and Benchmarks Track, 2025. 10.48550/arXiv.2503.13657.

[30] N.F. Liu, K. Lin, J. Hewitt, A. Paranjape, M. Bevilacqua, F. Petroni, P. Liang, Lost in the middle: how language models use long contexts, Trans. Assoc. Comput. Linguist. 12 (2024) 157–173. 10.1162/tacl_a_00638.

[31] L. Zheng, W.-L. Chiang, Y. Sheng, S. Zhuang, Z. Wu, Y. Zhuang, Z. Lin, Z. Li, D. Li, E.P. Xing, H. Zhang, J.E. Gonzalez, I. Stoica, Judging LLM-as-a-judge with MT-Bench and Chatbot Arena, in: Adv. Neural Inf. Process. Syst. 36 (NeurIPS 2023), Datasets and Benchmarks Track, 2023. 10.48550/arXiv.2306.05685.

[32] A. Panickssery, S.R. Bowman, S. Feng, LLM evaluators recognize and favor their own generations, in: Adv. Neural Inf. Process. Syst. 37 (NeurIPS 2024), 2024: pp. 68772–68802. 10.52202/079017-2197.

[33] T.A. Kass-Hout, Z. Xu, M. Mohebbi, H. Nelsen, A. Baker, J. Levine, E. Johanson, R.A. Bright, OpenFDA: an innovative platform providing access to a wealth of FDA’s publicly available data, J. Am. Med. Inform. Assoc. 23 (2016) 596–600. 10.1093/jamia/ocv153.

[34] Observational Health Data Sciences and Informatics, ATLAS and WebAPI. https://github.com/OHDSI/Atlas and https://github.com/OHDSI/WebAPI (accessed 29 October 2026).

[35] LangChain, LangGraph. https://github.com/langchain-ai/langgraph (accessed 29 October 2026).

[36] W. Kwon, Z. Li, S. Zhuang, Y. Sheng, L. Zheng, C.H. Yu, J.E. Gonzalez, H. Zhang, I. Stoica, Efficient memory management for large language model serving with PagedAttention, in: Proc. 29th Symp. Oper. Syst. Princ. (SOSP), ACM, 2023: pp. 611–626. 10.1145/3600006.3613165.

[37] J. Cohen, A coefficient of agreement for nominal scales, Educ. Psychol. Meas. 20 (1960) 37–46. 10.1177/001316446002000104.

[38] J. Gallifant, M. Afshar, S. Ameen, Y. Aphinyanaphongs, S. Chen, G. Cacciamani, D. Demner-Fushman, D. Dligach, R. Daneshjou, C. Fernandes, L.H. Hansen, A. Landman, L. Lehmann, L.G. McCoy, T. Miller, A. Moreno, N. Munch, D. Restrepo, G. Savova, R. Umeton, J.W. Gichoya, G.S. Collins, K.G.M. Moons, L.A. Celi, D.S. Bitterman, The TRIPOD-LLM reporting guideline for studies using large language models, Nat. Med. 31 (2025) 60–69. 10.1038/s41591-024-03425-5.

[39] J.R. Landis, G.G. Koch, The measurement of observer agreement for categorical data, Biometrics 33 (1977) 159–174. 10.2307/2529310.

