## Supplementary Meterials for "MENTOR: An Evidence-Informed Large Language Model-Based Multi-Agent System for Efficient Selection of Institution-Specific Negative Control Outcomes"

**S1. Hyperparameters and environment**

| **Item** | **Value** |
| --- | --- |
| LLM (all agents) | Qwen/Qwen3.6-27B-FP8, served with vLLM 0.23.0 (OpenAI-compatible API); maximum context length 32,768 tokens; prefix caching disabled |
| Hardware | 2 × NVIDIA RTX A6000 (48 GB each), tensor parallel |
| Temperature | 0 (all agents) |
| Reasoning (thinking) mode | Main: Adjudicator only; ablations: none or all agents |
| Maximum output tokens | Not set (limited by the 32,768-token context length) |
| Retries for malformed output | 3 (Suggester, Adjudicator, Diagnostician) |
| Concept hierarchy | Ancestors up to 3 levels (concept_ancestor), in Suggester and Adjudicator prompts |
| openFDA | Labels matched on substance or generic name; single-ingredient labels only (all labels if none); up to 1,000 labels per ingredient; most frequent version of each section; sections used: indications and usage, mechanism of action, contraindications, and four adverse-effect sections (boxed warning, adverse reactions, warnings and precautions, and warnings) |
| CEM | OHDSI WebAPI, public ATLAS instance, source key CEM; proposed = negativeControl or optimizedOut |
| Search (experiments) | min_patients = 10,000; floor = 10,000; step = 1,000 (single pass) |
| Search (default) | min_patients = 70,000; step 10,000; floor configurable |
| Diagnostician target | 100 NCOs (effective count) |
| Repetitions | 1 per configuration and drug pair (determinism verified, S14) |
| Orchestration | LangGraph 1.2.10, langchain-core 1.5.3, Python 3.10.14 |
| LLM judge | claude-opus-5 (Anthropic API), maximum 2,048 output tokens, default temperature, up to 5 retries |

CEM, Common Evidence Model; NCO, negative control outcome.

**S2. Prompts**

All prompts are sent as a single user message. Text in braces is replaced at run time: {drug_info} is the label text of one ingredient; {drug_infos} is the list of drug contexts; {outcome_name} is the candidate concept name; {hierarchy} is the list of ancestor concepts (or "(not provided)" when the hierarchy is disabled); {cem_verdict}, {cem_evidence}, and {first_judgment} are the CEM verdict, the evidence counts and flags, and the first-pass result and rationale; {target_nco_count}, {nco_count}, and {nco_list} are the target, the current number of NCOs, and their names.

**S2.1 Curator: drug label summarization**

```
You are an expert in clinical pharmacology.

[Task]
Summarize the drug information below concisely while preserving all clinically meaningful content.

[Drug information]
{drug_info}

[How to summarize]
1. Keep the JSON key structure EXACTLY as given. Do not add, remove, or rename keys.
2. Keep "ingredient_name" unchanged. Copy it verbatim.
3. For the other fields, condense the text into a compact list of distinct, standardized medical terms.
   - Remove duplicates, marketing language, dosage details, and study citations.
   - Use standard clinical terminology (prefer generic disease/event names).
   - Do NOT invent information that is not present in the input.
   - Do NOT drop any distinct condition or event; merge only true synonyms.
4. Each summarized field must be a single string, with items separated by "; ".
5. If a field is empty or unknown, output an empty string "".

[Output format]
- Output ONLY a single JSON object in exactly this format:
{"ingredient_name": "...", "indications": "...", "mechanism_of_action": "...", "contraindications": "...", "adverse_effects": "..."}
- Do NOT output any text outside the JSON object (no preamble, reasoning, or code fences).
```

**S2.2 Suggester: first-pass judgment**

You are an expert in pharmacoepidemiology.

[Task]
Judge whether the outcome below is a valid negative control outcome (NCO) for the given drug(s).

[Definition: negative control outcome (NCO)]
A negative control outcome is an outcome (disease, symptom, or event) that has NO causal relationship with any of the given drugs. NCOs are used to detect residual confounding and systematic bias in observational studies. An outcome is a valid NCO ONLY IF it meets ALL of the following for EVERY given drug:
1. It is NOT an indication of the drug (a condition the drug treats or prevents).
2. It is NOT a contraindication or a known adverse effect of the drug.
3. It has NO causal relationship through the drug's mechanism of action, metabolism, or pharmacokinetic/pharmacodynamic pathways, i.e. there is no biological basis for the drug to cause or prevent it.
If the outcome violates ANY of the three conditions, it is NOT a valid NCO.

[Drug information (reference only, not exhaustive)]
{drug_infos}

[Outcome]
{outcome_name}

[Concept hierarchy]
{hierarchy}

[How to reason]
- The drug information is REFERENCE ONLY. It is not an exhaustive or authoritative list.
- Do NOT conclude that there is no causal relationship simply because the outcome is absent from the drug information. Absence from the list is NOT evidence of no relationship.
- Base your judgment on your own expert knowledge of the drug's mechanism of action, metabolism, pharmacokinetics/pharmacodynamics, and the pathophysiology of the outcome.
- Reason from the biological mechanism linking (or not linking) the drug and the outcome, using the drug information only as supporting context.

[Decision]
- result = 1 : the outcome IS a valid NCO (it meets all three conditions).
- result = 0 : the outcome is NOT a valid NCO (it violates at least one condition).

[Output format]
- Output ONLY a single JSON object in exactly this format: {"result": 0, "reason": ""}
- "reason": 2-4 complete sentences. If result = 1, explain why there is no causal relationship, grounded in the drug's mechanism and the outcome's pathophysiology (not merely its absence from the drug information). If result = 0, name the violated condition and the specific relationship (indication, contraindication, adverse effect, or mechanistic pathway).
- Do NOT output any text outside the JSON object (no preamble, reasoning, or code fences).

**S2.3 Adjudicator: resolution of discordant candidates**

You are an expert in pharmacoepidemiology acting as an adjudicator.

[Task]
Two sources disagree on whether the outcome below is a valid negative control outcome (NCO) for the given drug(s):
- CEM (Common Evidence Model): an automated screen based on existing evidence of drug-outcome associations from publications, product labels, and spontaneous reports.
- A first-pass LLM judgment based on pharmacological reasoning (it may be unavailable if that step failed).
Resolve the disagreement and make the final decision.

[Definition: negative control outcome (NCO)]
A negative control outcome is an outcome (disease, symptom, or event) that has NO causal relationship with any of the given drugs. NCOs are used to detect residual confounding and systematic bias in observational studies. An outcome is a valid NCO ONLY IF it meets ALL of the following for EVERY given drug:
1. It is NOT an indication of the drug (a condition the drug treats or prevents).
2. It is NOT a contraindication or a known adverse effect of the drug.
3. It has NO causal relationship through the drug's mechanism of action, metabolism, or pharmacokinetic/pharmacodynamic pathways, i.e. there is no biological basis for the drug to cause or prevent it.
If the outcome violates ANY of the three conditions, it is NOT a valid NCO.

[Drug information (reference only, not exhaustive)]
{drug_infos}

[Outcome]
{outcome_name}

[Concept hierarchy]
{hierarchy}

[CEM verdict]
{cem_verdict}

[CEM evidence]
{cem_evidence}

[First-pass LLM judgment]
{first_judgment}

[How to read the CEM evidence]
Each number is a count of evidence items linking the drug and the outcome, at four matching levels:
- exact: the outcome concept itself; descendant: narrower (more specific) concepts; parent: the direct parent concept; ancestor: broader (more general) concepts.
Sources:
- Publications: drug-outcome relationships found in the published medical literature. A publication can describe any relationship, including the drug being studied as a treatment for the outcome, so it signals an association but not its direction.
- Product labels: the outcome is listed as an adverse reaction in official drug labels.
- Spontaneous reports: the drug-outcome pair shows a statistical signal in spontaneous adverse event reports. These signals are prone to reporting bias and confounding by indication: complications of the disease the drug treats can be flagged without being caused by the drug.
How CEM decides:
- ANY evidence from any source at any matching level excludes the outcome from the NCO list; the closer all counts are to 0, the better the candidate.
- This rule is deliberately strict. Evidence that exists only at the parent/ancestor level comes from broader concepts and may not apply to this specific outcome.
- Zero evidence does NOT prove there is no relationship; CEM coverage is incomplete.
Flags: indication_or_contraindication=1 means CEM matched an indication/contraindication; too_broad=1 means the concept is too general to be a useful NCO; drug_induced=1 means the concept itself implies drug causation; pregnancy_related=1 marks pregnancy outcomes; not_prevalent=1 means the outcome is rare; optimized_out=1 means CEM dropped it as redundant with a related concept.

[How to decide]
- The drug information is REFERENCE ONLY. It is not an exhaustive or authoritative list.
- Do NOT conclude that there is no causal relationship simply because the outcome is absent from the drug information. Absence from the list is NOT evidence of no relationship.
- Base your judgment on your own expert knowledge of the drug's mechanism of action, metabolism, pharmacokinetics/pharmacodynamics, and the pathophysiology of the outcome.
- Then apply the following rules:
1. Identify which source is likely wrong and why. Do not side with either source by default.
2. An exact or descendant product label count means a labeled adverse reaction (condition 2 is violated) unless the label match is clearly a different condition.
3. Other exact/descendant evidence (publications, spontaneous reports) combined with a plausible pathway (indication, adverse effect, or mechanism) means the outcome is NOT a valid NCO.
4. If the only evidence is parent/ancestor-level, or is explainable by confounding by indication, and you find no mechanistic link, the outcome can be a valid NCO.
5. If you remain uncertain after weighing everything, choose result = 0. A false NCO biases calibration more than a missing one.

[Decision]
- result = 1 : the outcome IS a valid NCO (it meets all three conditions).
- result = 0 : the outcome is NOT a valid NCO (it violates at least one condition).

[Output format]
- Output ONLY a single JSON object in exactly this format: {"result": 0, "reason": ""}
- "reason": 2-4 complete sentences. State which source you sided with, the decisive evidence or mechanism, and why the other source was wrong.
- Do NOT output any text outside the JSON object (no preamble, reasoning, or code fences).

**S2.4 Diagnostician: sufficiency of the NCO set**

You are an expert in pharmacoepidemiology.

[Task]
Judge whether the current set of negative control outcomes (NCOs) for the given drug(s) is sufficient for the study, or whether the search should continue with a lower patient-count threshold to find more candidates.

[Definition: negative control outcome (NCO)]
A negative control outcome is an outcome (disease, symptom, or event) that has NO causal relationship with any of the given drugs. NCOs are used to detect residual confounding and systematic bias in observational studies. An outcome is a valid NCO ONLY IF it meets ALL of the following for EVERY given drug:
1. It is NOT an indication of the drug (a condition the drug treats or prevents).
2. It is NOT a contraindication or a known adverse effect of the drug.
3. It has NO causal relationship through the drug's mechanism of action, metabolism, or pharmacokinetic/pharmacodynamic pathways, i.e. there is no biological basis for the drug to cause or prevent it.
If the outcome violates ANY of the three conditions, it is NOT a valid NCO.

[Target]
Target number of NCOs: {target_nco_count}

[Drug information (reference only, not exhaustive)]
{drug_infos}

[NCO set]
Raw number of NCOs: {nco_count}
One outcome name per line.
{nco_list}

[How the set was built]
Each outcome above was validated individually against the definition, and every outcome that passed is listed. The set has NOT been optimized as a whole:
- Hierarchically related concepts are all kept. If a parent concept and its child concepts are all valid, they are listed separately (e.g. a general condition and several of its specific subtypes).
- Near-synonymous or heavily overlapping concepts may also be listed separately.
Such concepts largely capture the same patients and the same bias, so they count as roughly one NCO, not several. The raw number therefore overstates the number of independent NCOs.

[How to judge]
1. Effective count: estimate how many independent NCOs remain after collapsing parent/child and near-synonymous concepts into one. Compare this effective count, not the raw number, with the target.
2. Diversity: the set should span many organ systems, disease types, and clinical domains. A set dominated by one body region or disease category cannot detect bias broadly, even if it is large.
3. Do NOT re-judge whether individual outcomes are valid NCOs; the drug information is context for judging diversity only.

[Decision]
- result = 1 : sufficient. The effective count is at or above the target, and diversity is adequate.
- result = 0 : insufficient. The effective count is below the target, or the set is concentrated in a few categories; continue the search.

[Output format]
- Output ONLY a single JSON object in exactly this format: {"result": 0, "reason": ""}
- "reason": 2-4 complete sentences. State the raw and estimated effective counts relative to the target, and which categories are well covered and which are missing or overrepresented.
- Do NOT output any text outside the JSON object (no preamble, reasoning, or code fences).

**S2.5 LLM judge (reference standard; blinded)**

You are an expert in pharmacoepidemiology acting as an independent reviewer.

[Task]
Decide whether the outcome below is a valid negative control outcome (NCO) for the given drug(s). Your decision is used as a reference label for evaluating an automated NCO selection system.

[Definition: negative control outcome (NCO)]
A negative control outcome is an outcome (disease, symptom, or event) that has NO causal relationship with any of the given drugs. NCOs are used to detect residual confounding and systematic bias in observational studies. An outcome is a valid NCO ONLY IF it meets ALL of the following for EVERY given drug:
1. It is NOT an indication of the drug (a condition the drug treats or prevents).
2. It is NOT a contraindication or a known adverse effect of the drug.
3. It has NO causal relationship through the drug's mechanism of action, metabolism, or pharmacokinetic/pharmacodynamic pathways, i.e. there is no biological basis for the drug to cause or prevent it.
If the outcome violates ANY of the three conditions, it is NOT a valid NCO.

[Outcome]
{outcome_name}

[Drug information (reference only, not exhaustive)]
{drug_infos}

[How to judge]
- Check each of the three conditions for every given drug in turn.
- The drug information is a summary of the drug labels and may be incomplete. An outcome missing from it is not, by that fact alone, unrelated to the drug.
- Draw on your own knowledge of the drugs' pharmacology and of the outcome's pathophysiology and epidemiology.

[Decision]
- result = 1 : the outcome IS a valid NCO (it meets all three conditions).
- result = 0 : the outcome is NOT a valid NCO (it violates at least one condition).

[Output format]
- Output ONLY a single JSON object in exactly this format: {"result": 0, "reason": ""}
- "reason": 2-4 complete sentences. Name the deciding condition and the specific drug-outcome relationship (for result = 0), or explain why no relationship exists for any of the given drugs (for result = 1).
- Do NOT output any text outside the JSON object (no preamble, reasoning, or code fences).

Example of the {hierarchy} input:

Parent concepts (up to 3 levels, nearest first):
- Disorder of function of stomach (level 1)
- Indigestion (level 1)
- Dyspepsia (level 2)
- Disorder of digestive organ (level 3)
- Gastric disorder (level 3)

Example of the {cem_evidence} input:

Evidence counts (exact / descendant / parent / ancestor):
- Publications: 0 / 0 / 1 / 5
- Product labels: 0 / 0 / 0 / 0
- Spontaneous reports: 0 / 0 / 0 / 0
Flags: indication_or_contraindication=0, too_broad=0, drug_induced=0, pregnancy_related=0, not_prevalent=0, optimized_out=0

**S3. Adjudicator inputs**

For each discordant candidate, the Adjudicator receives the drug context, the candidate concept name and its ancestor concepts, the CEM verdict, the CEM evidence counts by source and matching level with the CEM flags, and the first-pass judgment and rationale. The decision rules are those listed in Section 3.1.4 and in the adjudicator prompt (S2.3). Examples of the {hierarchy} and {cem_evidence} inputs are shown at the end of S2.

**S4. Data and knowledge-base versions**

| **Resource** | **Version / date** |
| --- | --- |
| OMOP CDM | Version 5.3 (AUSOM; visits from January 1994 to November 2025) |
| OMOP vocabulary | v5.0 27-FEB-25 (SNOMED CT International 2024-08-01; RxNorm 2024-12-02; RxNorm Extension 2024-12-15) |
| AUSOM | 2,938,928 persons |
| CEM | Public ATLAS instance (WebAPI 2.14.0; CEM source vocabulary v5.0 29-FEB-24), queried October 2026; CEM data release not reported by the API |
| openFDA drug label | Queried October 2026 |

**S5. Construction of the evaluation set**

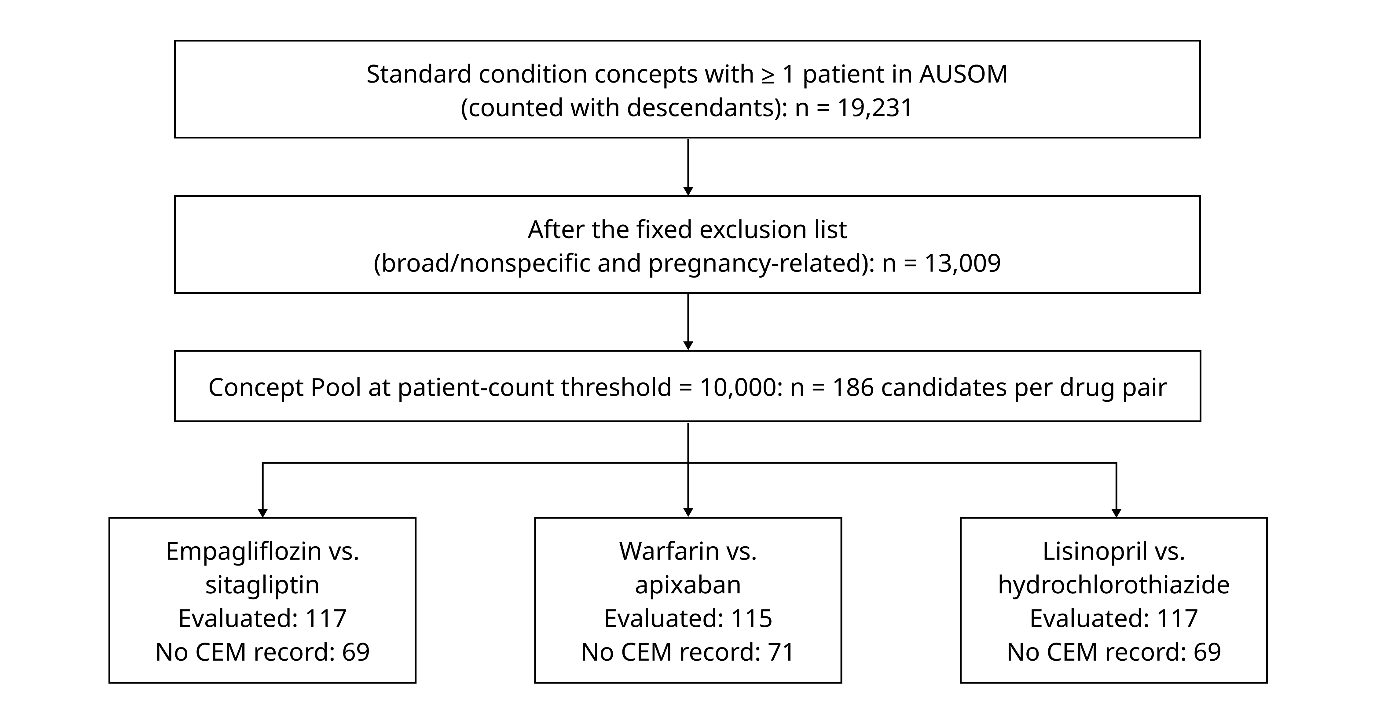

Flow of candidate selection for the evaluation set. AUSOM, Ajou University School of Medicine; CEM, Common Evidence Model.

Construction of the evaluation set. Candidates are drawn from the institution's Concept Pool, which is independent of the drug pair; candidates without a CEM record for a pair are outside the system's scope.

Exclusion list. The Concept Pool is built separately for each institution, whereas the exclusion list is fixed and shared across institutions. The exclusion list consists of two groups of concepts that are unsuitable as NCOs regardless of the drug pair. The first group comprises overly broad or nonspecific concepts and was constructed from the OMOP vocabulary using four rules: (i) descendants of two high-level condition concepts (concept IDs 40481841 and 4147183); (ii) concepts having more than 45 standard condition descendants, indicating a high-level node in the hierarchy; (iii) concepts directly related to concept 4008453 through the concept relationship table; and (iv) concepts whose names contain generic terms such as "finding," "disorder of," "disease of," "injury," "symptoms," "abnormal," "by site," "by mechanism," or "of body region." A curated list of approximately 80 additional nonspecific concepts (e.g., malaise, generally unwell, problem, illness, complication of medical care) was appended. The second group comprises pregnancy-related concepts, defined as all descendants of 12 pregnancy-related ancestor concepts plus three individually specified concepts, which were excluded because their occurrence is governed by reproductive status rather than drug exposure. In total, the exclusion list contained 15,951 concepts.

**S6. Code**

Code repository: https://github.com/ABMI/MENTOR.

Graph definition. The system is implemented as a LangGraph StateGraph over a shared state that holds the input ingredient concepts and their drug context, the current patient-count threshold (min_patients), and every candidate outcome with its patient count, CEM verdict and evidence, first-pass judgment, Adjudicator judgment, and final decision. The top-level graph is START → Curator → Suggester → Diagnostician. The Curator routes to END if any input is not a valid ingredient concept. The Diagnostician routes back to the Suggester with min_patients lowered by a fixed step (10,000 by default) unless the NCO set is judged sufficient or the next threshold would fall below a configurable floor, in which case the run ends. Candidates added in earlier iterations keep their decisions and are not reviewed again.

Running the system. Installation and usage instructions are provided in the repository. Each run returns the NCO list with rationales, an ATLAS concept set expression, and a CohortGenerator negative control outcome table.

**S7. Human sample design**

| **Drug pair** | **Evaluation set, n** | **Sample, n** | **Valid** | **Not valid** | **Indeterminate** |
| --- | --- | --- | --- | --- | --- |
| Empagliflozin vs. sitagliptin | 117 | 30 | 13 | 11 | 6 |
| Warfarin vs. apixaban | 115 | 30 | 16 | 1 | 13 |
| Lisinopril vs. HCTZ | 117 | 30 | 20 | 2 | 8 |
| Total | 349 | 90 | 49 | 14 | 27 |

Stratified random sampling with proportional allocation; one epidemiologist with 7–10 years of experience reviewed each drug pair, blinded to the CEM, MENTOR, and judge labels. CEM, Common Evidence Model; HCTZ, hydrochlorothiazide.

Labeling guideline (summary). Reviewers judged, for each drug pair, whether each outcome is a valid NCO, i.e., whether there is credible evidence that either drug causes or prevents it. An outcome was not a valid NCO if it is an indication (including complications of the indication), a known adverse effect, an expected pharmacological effect, or a causal association reported in the literature, regardless of direction; reviewers were asked to distinguish causal relationships from associations due to confounding. Labels were valid NCO (1), not a valid NCO (0), or indeterminate (99), with optional confidence (high, moderate, low) and a one- to two-line reason. Permitted sources were pharmacological knowledge, product labels and prescribing information, peer-reviewed literature, and drug information databases; prohibited were the use of generative AI for labeling decisions, information on which method proposed a candidate, the outputs of the evaluated tools, and consultation with other reviewers. Reviewers were asked to form a preliminary judgment from their own knowledge before consulting sources. Generative AI could be used only to translate or summarize literature, and any such use was logged (model, question, item).

**S8. Error coding sheet** (all FPs and FNs of the main configuration with case records and LLM-assigned codes) — error_coding_sheet.xlsx.

**S9. Error-analysis codebook**

```
Unit of analysis: one misclassified candidate of the main configuration, relative to the blinded LLM judge.
  FP (false positive): the system accepted the outcome as a valid NCO; the judge rejected it.
  FN (false negative): the system rejected the outcome; the judge accepted it.
Decision path (assigned automatically):
  concordant: CEM and the first-pass (Suggester) judgment agreed; the shared decision was final. The deciding rationale is the Suggester's.
  adjudicated: CEM and the first pass disagreed; the Adjudicator decided. The deciding rationale is the Adjudicator's.

A. Error type (FP only) - the nature of the evidence that invalidates the outcome, judged against the drug context the system was given:
  Type 1 (label-grounded, direct): the outcome name or a synonym appears verbatim in the drug context.
  Type 2 (label-grounded, indirect): the outcome is not named, but is reachable from a drug-context item through a chain of established
          clinical relationships (a broader or narrower category, a defining sign or symptom, or downstream consequences of a listed effect),
          regardless of the number of steps.
  Type 3 (off-label, established): absent from the drug context and not reachable from it, but supported by established clinical or
          epidemiological evidence.
  Type 4 (speculative): the evidence is limited to surrogate markers, preclinical findings, or class effects of other drugs, or the chain
          from the drug context contains a speculative link.
  If several apply, choose the one most directly responsible for invalidation (lowest number that holds).
  Context anchor: quote the drug-context term(s) used for Type 1 or Type 2.
  Inference steps: the number of inference steps from the context anchor to the outcome (0 for Type 1; 1, 2, ... for Type 2;
          0 for Type 3 and Type 4). Example: listed syncope -> fall -> rib fracture = 2 steps.

B. Failure mode (FP only) - what the deciding agent's rationale did wrong.
  First decide whether the rationale RECOGNIZED the relevant drug-context term (named it, or the listed effect it leads to):
  - Recognized, but the outcome was still accepted:
    label_dismissed: named the listed adverse effect (or its broader/narrower term) but accepted the outcome anyway, arguing that the
                     listing is not causal (e.g., background incidence in trials, no mechanism). Under the NCO definition a known
                     adverse effect invalidates the outcome regardless of mechanism.
    label_override: (adjudicated only) recognized the listed term but accepted the outcome because the CEM evidence was absent or
                    only at the parent/ancestor level.
    no_downstream: acknowledged a listed effect but rejected the causal chain (one or more steps) from it to the outcome.
  - Not recognized (the rationale does not name the relevant context term):
    taxonomic_mismatch: failed to link the outcome to a broader, narrower, or synonymous term listed in the context
                        (e.g., states that the outcome is not a known adverse effect although its parent term is listed).
    mechanism_first: excluded a relationship from the drug's primary mechanism without checking the adverse effects listed in the context.
    knowledge_gap: the invalidating evidence is not in the context and the rationale shows no awareness of it (typical of Type 3/4).
  other: none of the above; explain in the note.
  Also record whether the relevant broader term appeared in the [Concept hierarchy] input (hierarchy_had_anchor: yes/no/n.a.).

C. FN category (FN only) - why the deciding agent rejected a valid outcome:
  over_extended_mechanism: rejected through an indirect, weak, or speculative pathway (e.g. bleeding risk -> gastritis).
  indication_confusion: treated a comorbidity or complication of the indication (or the indication's disease course) as a causal relationship.
  cem_overweighted: (adjudicated, or CEM-driven) rejected mainly because CEM reported evidence, without a plausible pathway.
  label_misreading: matched the outcome to a drug-context item that is a different condition.
  other: none of the above; explain in the note.
```

**S10. TRIPOD-LLM checklist**

Study design: LLM evaluation (with a methods component: an LLM-based multi-agent system without model training); task: classification. Item topics are abbreviated; see Gallifant et al., Nat Med 2025;31:60–69 for the full wording. N/A, not applicable to this design or task.

| **Item** | **Topic** | **Reported in** |
| --- | --- | --- |
| 1 | Title: LLM development/evaluation, task, population, outcome | Title |
| 2 | Abstract (TRIPOD-LLM for Abstracts) | Abstract |
| 3a | Background: healthcare context, use case, rationale | Introduction, paragraphs 1–2; Section 2 |
| 3b | Target population, intended use, intended users | Introduction, paragraph 3; Discussion, paragraphs 3–5 |
| 4 | Objectives, including development/validation stages | Introduction, paragraph 3 |
| 5a | Data sources and rationale | Methods 3.2.1–3.2.3 |
| 5b | Data points and their distribution | Results (candidate pool and human sample); S7; S5 |
| 5c | Dates of the oldest and newest text | Methods 3.2.2 (CEM queried October 2026); S4 (openFDA queried October 2026) |
| 5d | Pre-processing and quality checks | Methods 3.1.2 (label selection and summarization), 3.2.3 (exclusion list) |
| 5e | Missing/imbalanced data, exclusions | Methods 3.1.3, 3.2.3 (no CEM record excluded); Results; S1 (retries) |
| 6a | LLM name, version, training cut-off | Methods 3.2.4; S1; training data cut-off not reported by the developer (model released April 2026) |
| 6b | Development details (architecture, training, fine-tuning) | Methods 3.1.1–3.1.5 (system architecture); no training or fine-tuning |
| 6c | Text generation details: prompting, inference settings | Methods 3.1.2–3.1.5, 3.2.4; S2; S1 |
| 6d | Initial and post-processed output format | Methods 3.1.3; S2 (JSON result and reason) |
| 6e | Classification rationale, probabilities, thresholds | Methods 3.1.3, 3.2.6 (binary output; no probability threshold) |
| 7a | Quality metrics for generated text, error types | N/A (classification); rationales examined in the error analysis (Methods 3.2.7d) |
| 7b | Metric relevance; agreement with human evaluation | Methods 3.2.5–3.2.6; Results (judge validation, human-label consistency); S11; S12 |
| 7c | Outcome definition, inference dates, metrics | Methods 3.1.3 (NCO definition), 3.2.4, 3.2.6 |
| 7d | Assessor qualifications, instructions, inter-assessor agreement | Methods 3.2.5 (three epidemiologists, 7–10 years of experience); S7 (guideline); one reviewer per pair, agreement not assessed (Discussion, limitations) |
| 7e | Comparison with other LLMs, humans, benchmarks | Methods 3.2.2 (CEM), 3.2.7a (component analysis); Table 1; Figure 2; S15 |
| 8a | Labeling method and annotation guidelines | Methods 3.2.5; S7 |
| 8b | Number of annotators, double annotation, agreement | Methods 3.2.5 (three reviewers, one per pair; no double annotation) |
| 8c | Annotator background; characteristics of labeling models | Methods 3.2.5 (LLM judge; three epidemiologists), 3.2.7d (error coding model) |
| 9a | Prompt design, curation, selection | Methods 3.1.2–3.1.5, 3.2.4; S2 |
| 9b | Data used to develop prompts | Methods 3.2.4 (study drug pairs); Discussion, limitations |
| 10 | Pre-processing before summarization | Methods 3.1.2 (Curator label summarization) |
| 11 | Instruction tuning/alignment | N/A |
| 12 | Compute requirements | Methods 3.2.4; Results (adjudication load and cost); S1; S19 |
| 13 | Ethical approval and consent | Methods 3.2.8 (AJOUIRB-EX-2026-412) |
| 14a | Funding and role of funder | Funding |
| 14b | Conflicts of interest | Declaration of Competing Interest |
| 14c | Protocol | N/A (no protocol prepared) |
| 14d | Registration | N/A (not registered) |
| 14e | Data availability | Data Availability; Supplementary Data |
| 14f | Code availability | Code Availability; S6 |
| 15 | Patient and public involvement | N/A (no patient or public involvement) |
| 16a | Flow of data | Results; S5 |
| 16b | Characteristics by data source and split | Results (candidate pool); S7 |
| 16c | Development vs. evaluation data | N/A (no training data); prompts developed on the study drug pairs (Methods 3.2.4) |
| 16d | Counts per analysis | Table 1; S11; S15; S18 |
| 17 | Performance on pre-specified metrics, human evaluation | Results; Table 1; Figure 2; S15 |
| 18 | LLM updating | N/A |
| 19a | Overall interpretation, including fairness | Discussion, paragraphs 1–2 (fairness not assessed) |
| 19b | Limitations | Discussion, paragraphs 8–9 |
| 19c | Known data challenges | Discussion, paragraphs 4, 7, and 9 |
| 19d | Intended use: input, end user, oversight | Introduction, paragraph 3; Discussion, paragraph 3 |
| 19e | Handling of poor-quality or unavailable input | Methods 3.1.2–3.1.3 (ingredient validation, retries, out-of-scope candidates) |
| 19f | Required user interaction and expertise | Discussion, paragraphs 3 and 5 |
| 19g | Future research | Section 6 (Conclusion) |

**S11. Agreement of the judge with expert review**

**(a) Agreement statistics**

| **Analysis** | **n** | **Accuracy** | **κ** | **PPV** | **NPV** | **Sensitivity** | **Specificity** |
| --- | --- | --- | --- | --- | --- | --- | --- |
| Primary: indeterminate excluded | 63 | 0.86  (0.76–0.94) | 0.63  (0.39–0.83) | 0.95  (0.89–1.00) | 0.63  (0.41–0.86) | 0.86  (0.75–0.95) | 0.86  (0.64–1.00) |
| *By drug pair* | | | | | | | |
| Empagliflozin vs. sitagliptin | 24 | 0.88  (0.75–1.00) | 0.75  (0.47–1.00) | 0.86  (0.67–1.00) | 0.90  (0.67–1.00) | 0.92  (0.75–1.00) | 0.82  (0.56–1.00) |
| Warfarin vs. apixaban | 17 | 0.88  (0.71–1.00) | 0.45  (0.00–1.00) | 1.00  (1.00–1.00) | 0.33  (0.00–1.00) | 0.88  (0.69–1.00) | 1.00  (1.00–1.00) |
| Lisinopril vs. HCTZ | 22 | 0.82  (0.64–0.95) | 0.42  (0.00–0.83) | 1.00  (1.00–1.00) | 0.33  (0.00–0.75) | 0.80  (0.60–0.95) | 1.00  (1.00–1.00) |
| *Sensitivity analyses (indeterminate ratings reassigned)* | | | | | | | |
| Indeterminate as not valid | 90 | 0.70  (0.60–0.79) | 0.38  (0.18–0.56) | 0.68  (0.56–0.79) | 0.75  (0.57–0.90) | 0.86  (0.75–0.94) | 0.51  (0.36–0.67) |
| Indeterminate as valid | 90 | 0.80  (0.72–0.88) | 0.46  (0.24–0.65) | 0.97  (0.92–1.00) | 0.43  (0.24–0.62) | 0.79  (0.70–0.88) | 0.86  (0.64–1.00) |

Judge sensitivity, specificity, PPV, and NPV are computed with the expert label as the reference and a valid NCO as the positive class. Values are point estimates with bootstrap 95% CIs. Per-pair rows use the primary analysis; each pair was reviewed by one expert. Disagreements: judge not valid/expert valid, 7; judge valid/expert not valid, 2 (exact McNemar P = 0.18). CI, confidence interval; HCTZ, hydrochlorothiazide; NPV, negative predictive value; PPV, positive predictive value; κ, Cohen’s kappa.

**(b) Confusion matrix**

|  | **Expert: valid** | **Expert: not valid** | **Expert: indeterminate** |
| --- | --- | --- | --- |
| Judge: valid | 42 | 2 | 18 |
| Judge: not valid | 7 | 12 | 9 |

**S12. Disagreements between the judge and the experts**

| **Drug pair** | **Outcome** | **Judge** | **Expert** | **Judge rationale (abridged)** |
| --- | --- | --- | --- | --- |
| Empagliflozin vs. sitagliptin | Late effects of respiratory tuberculosis | valid | not valid | Late effects (sequelae) of respiratory tuberculosis are fibrotic/structural residua of a prior mycobacterial infection, and neither empagliflozin (SGLT2 inhibition with glycosuria and natriuresis) nor sitagliptin (DPP-4 inhibition increasing incretins) treats or prevents this condition, so it is not an indication. |
| Empagliflozin vs. sitagliptin | Chronic hepatitis | valid | not valid | Chronic hepatitis (predominantly chronic viral hepatitis B/C or autoimmune hepatitis) is neither an indication nor a contraindication for empagliflozin or sitagliptin, and neither SGLT2 inhibition nor DPP-4 inhibition has a plausible mechanism for causing or preventing a chronic hepatitic process. |
| Empagliflozin vs. sitagliptin | Gastrointestinal mucositis | not valid | valid | Condition 2 is violated for sitagliptin: its labeling lists mouth ulceration and stomatitis as adverse reactions, and DPP-4 inhibitors are also linked to mucosal involvement in drug-induced bullous pemphigoid, providing a recognized mechanism for inflammatory/ulcerative lesions of the gastrointestinal (including oral) mucosa. |
| Lisinopril vs. HCTZ | Subdural hemorrhage | not valid | valid | Subdural hemorrhage is not an indication of lisinopril or hydrochlorothiazide, but condition 3 is violated because plausible pharmacologic pathways link both drugs to this outcome. |
| Lisinopril vs. HCTZ | Peripheral vertigo | not valid | valid | Peripheral vertigo (e.g., Ménière's disease, BPPV) fails conditions 1 and 2. Hydrochlorothiazide is a long-standing (off-label but guideline-recognized) treatment for Ménière's disease, a classic peripheral vestibular disorder, so the outcome is effectively an indication/preventable condition for that drug, and vertigo and dizziness are explicitly listed labeled adverse effects of hydrochlorothiazide. |
| Lisinopril vs. HCTZ | Postmenopausal osteoporosis | not valid | valid | Condition 3 is violated for hydrochlorothiazide: thiazide diuretics reduce renal calcium excretion by enhancing distal tubular calcium reabsorption, producing a well-documented increase in bone mineral density and a reduced risk of osteoporosis and osteoporotic fracture. |
| Lisinopril vs. HCTZ | Chronic hepatitis | not valid | valid | Both drugs have documented hepatotoxic potential, so hepatitis is not causally independent of them. Lisinopril labeling lists hepatic failure, and ACE inhibitors (including lisinopril) are reported to cause hepatocellular/cholestatic hepatitis that can be prolonged or progressive in case reports; hydrochlorothiazide lists intrahepatic cholestatic jaundice and hepatic coma. |
| Warfarin vs. apixaban | Postmenopausal osteoporosis | not valid | valid | Warfarin inhibits vitamin K epoxide reductase, which impairs gamma-carboxylation of vitamin K-dependent bone proteins such as osteocalcin and matrix Gla protein, providing a direct mechanistic basis for reduced bone mineral density and osteoporosis; observational studies comparing warfarin with DOACs (including apixaban) have specifically examined osteoporosis and osteoporotic fracture as effect outcomes. |
| Warfarin vs. apixaban | Chronic kidney disease stage 5 | not valid | valid | Warfarin has a plausible causal pathway to advanced chronic kidney disease: warfarin-related nephropathy (glomerular hemorrhage with red cell cast tubular obstruction) causes acute kidney injury—listed among its adverse effects—and accelerated progression of CKD, and warfarin-induced vascular/calciphylaxis-type calcification via inhibition of vitamin K-dependent matrix Gla protein further promotes renal decline. |

Candidates rated indeterminate by the experts are not listed. HCTZ, hydrochlorothiazide.

**S13. Candidate-level results for all drug pairs and configurations**
— Supplementary_Data.xlsx, sheet S13.

**S14. Reproducibility**

Two independent runs (main configuration and Adjudicator without reasoning) share identical Curator and Suggester settings. They produced identical drug contexts for all six ingredients and identical first-pass results and rationales (character for character) for all 349 candidates; the drug contexts were also identical in the run without the concept hierarchy. Repeated execution of the remaining configurations was therefore not performed.

**S15. Performance of all strategies and configurations, pooled and by drug pair (judge reference)**

**(a) Discordance-handling strategies and configuration ablations, pooled (n = 349)**

| **Configuration** | **NCOs** | **Accuracy** | **PPV** | **Sens.** | **Spec.** | **Δ Accuracy vs. main (95% CI)** | **P** | **Tokens (M)** | **Time (min)** |
| --- | --- | --- | --- | --- | --- | --- | --- | --- | --- |
| Discordance handling | | | | | | | | | |
| CEM only | 121 | 0.53 | 0.79 | 0.41 | 0.78 | −0.298  (−0.355, −0.241) | <0.001 | 0 | 3 |
| First-pass LLM only | 239 | 0.81 | 0.85 | 0.87 | 0.69 | −0.017  (−0.037, 0.000) | 0.11 | 0.65 | 25 |
| Adjudicator, no reasoning | 216 | 0.78 | 0.86 | 0.80 | 0.74 | −0.049  (−0.077, −0.020) | 0.002 | 1.06 | 36 |
| Adjudicator, reasoning (main) | 243 | 0.83 | 0.86 | 0.89 | 0.70 | Reference |  | 1.36 | 158 |
| Configuration ablations (full pipeline) | | | | | | | | | |
| Reasoning in all agents | 258 | 0.88 | 0.87 | 0.97 | 0.72 | 0.054  (0.017, 0.092) | 0.008 | 2.14 | 410 |
| Curator: ingredient names only | 231 | 0.81 | 0.86 | 0.85 | 0.72 | −0.023  (−0.057, 0.009) | 0.23 | 0.98 | 161 |
| Curator: raw label text | 250 | 0.83 | 0.85 | 0.91 | 0.67 | 0.003  (−0.029, 0.034) | >0.99 | 4.62 | 160 |
| No concept hierarchy | 246 | 0.81 | 0.84 | 0.88 | 0.66 | −0.020  (−0.049, 0.009) | 0.23 | 1.26 | 169 |

Discordance-handling strategies are computed from the same runs, because the CEM verdict and the first-pass judgment do not depend on the Adjudicator; the Adjudicator without reasoning is the run in which only the Adjudicator's reasoning mode was disabled, with identical first-pass judgments. Δ, configuration minus main; 95% CIs by bootstrap stratified by drug pair; P, exact McNemar test. Tokens and time are summed over the three drug pairs; for discordance-handling strategies, only the steps each strategy requires are counted (Diagnostician excluded), and for ablations the full run is counted. Time includes CEM waiting (about 2–3 minutes per configuration). CEM, Common Evidence Model; CI, confidence interval; LLM, large language model; M, million; NCO, negative control outcome; PPV, positive predictive value; Sens., sensitivity; Spec., specificity.

**(b) All configurations by drug pair**

| **Drug pair** | **Configuration** | **NCOs** | **Accuracy** | **PPV** | **NPV** | **Sens.** | **Spec.** |
| --- | --- | --- | --- | --- | --- | --- | --- |
| Empagliflozin vs. sitagliptin | CEM | 74 | 0.68 | 0.72 | 0.63 | 0.77 | 0.56 |
|  | First-pass LLM only | 84 | 0.80 | 0.77 | 0.88 | 0.94 | 0.60 |
|  | Main | 86 | 0.80 | 0.77 | 0.90 | 0.96 | 0.58 |
|  | Adjudicator, no reasoning | 83 | 0.81 | 0.78 | 0.88 | 0.94 | 0.62 |
|  | Reasoning in all agents | 81 | 0.86 | 0.83 | 0.94 | 0.97 | 0.71 |
|  | Curator: ingredient names only | 84 | 0.77 | 0.75 | 0.82 | 0.91 | 0.56 |
|  | Curator: raw label text | 89 | 0.79 | 0.75 | 0.93 | 0.97 | 0.54 |
|  | No concept hierarchy | 89 | 0.79 | 0.75 | 0.93 | 0.97 | 0.54 |
| Warfarin vs. apixaban | CEM | 30 | 0.44 | 0.93 | 0.27 | 0.31 | 0.92 |
|  | First-pass LLM only | 84 | 0.86 | 0.94 | 0.65 | 0.88 | 0.80 |
|  | Main | 87 | 0.90 | 0.95 | 0.75 | 0.92 | 0.84 |
|  | Adjudicator, no reasoning | 78 | 0.83 | 0.95 | 0.57 | 0.82 | 0.84 |
|  | Reasoning in all agents | 100 | 0.91 | 0.90 | 1.00 | 1.00 | 0.60 |
|  | Curator: ingredient names only | 79 | 0.85 | 0.96 | 0.61 | 0.84 | 0.88 |
|  | Curator: raw label text | 85 | 0.85 | 0.93 | 0.63 | 0.88 | 0.76 |
|  | No concept hierarchy | 87 | 0.85 | 0.92 | 0.64 | 0.89 | 0.72 |
| Lisinopril vs. HCTZ | CEM | 17 | 0.46 | 0.82 | 0.40 | 0.19 | 0.93 |
|  | First-pass LLM only | 71 | 0.77 | 0.83 | 0.67 | 0.80 | 0.72 |
|  | Main | 70 | 0.78 | 0.84 | 0.68 | 0.80 | 0.74 |
|  | Adjudicator, no reasoning | 55 | 0.70 | 0.85 | 0.56 | 0.64 | 0.81 |
|  | Reasoning in all agents | 77 | 0.87 | 0.88 | 0.85 | 0.92 | 0.79 |
|  | Curator: ingredient names only | 68 | 0.79 | 0.87 | 0.69 | 0.80 | 0.79 |
|  | Curator: raw label text | 76 | 0.85 | 0.87 | 0.80 | 0.89 | 0.77 |
|  | No concept hierarchy | 70 | 0.78 | 0.84 | 0.68 | 0.80 | 0.74 |

CEM, Common Evidence Model; HCTZ, hydrochlorothiazide; NCO, negative control outcome; NPV, negative predictive value; PPV, positive predictive value; Sens., sensitivity; Spec., specificity.

**(c) Differences from the main configuration in other metrics, pooled (n = 349)**

| **Configuration** | **Δ PPV (95% CI)** | **Δ NPV (95% CI)** | **Δ Sens. (95% CI)** | **Δ Spec. (95% CI)** |
| --- | --- | --- | --- | --- |
| Discordance handling | | | | |
| CEM only | −0.071  (−0.127, −0.019) | −0.369  (−0.441, −0.299) | −0.485  (−0.548, −0.421) | 0.078  (0.000, 0.153) |
| First-pass LLM only | −0.007  (−0.022, 0.009) | −0.037  (−0.072, −0.009) | −0.021  (−0.042, −0.004) | −0.009  (−0.045, 0.028) |
| Adjudicator, no reasoning | 0.005  (−0.011, 0.024) | −0.118  (−0.169, −0.066) | −0.094  (−0.131, −0.058) | 0.043  (0.009, 0.085) |
| Adjudicator, reasoning (main) | Reference | Reference | Reference | Reference |
| Configuration ablations (full pipeline) | | | | |
| Reasoning in all agents | 0.016  (−0.017, 0.050) | 0.148  (0.072, 0.227) | 0.073  (0.035, 0.113) | 0.017  (−0.064, 0.100) |
| Curator: ingredient names only | 0.001  (−0.028, 0.031) | −0.061  (−0.118, −0.004) | −0.043  (−0.079, −0.009) | 0.017  (−0.051, 0.090) |
| Curator: raw label text | −0.008  (−0.034, 0.016) | 0.024  (−0.040, 0.087) | 0.017  (−0.017, 0.053) | −0.026  (−0.091, 0.033) |
| No concept hierarchy | −0.019  (−0.042, 0.004) | −0.026  (−0.080, 0.027) | −0.009  (−0.040, 0.022) | −0.043  (−0.101, 0.010) |

Δ, configuration minus main; 95% CIs by bootstrap stratified by drug pair. Rows and the main configuration as in panel (a). PPV, positive predictive value; NPV, negative predictive value; Sens., sensitivity; Spec., specificity.

**(d) Accuracy on discordant candidates in the main configuration (n = 154)**

| **Configuration** | **All discordant (n = 154)** | **CEM not proposed, first pass valid (n = 136)** | **CEM proposed, first pass not valid (n = 18)** |
| --- | --- | --- | --- |
| CEM only | 0.18  (28/154) | 0.14  (19/136) | 0.50  (9/18) |
| First-pass LLM only | 0.82  (126/154) | 0.86  (117/136) | 0.50  (9/18) |
| Adjudicator, no reasoning | 0.75  (115/154) | 0.77  (105/136) | 0.56  (10/18) |
| Adjudicator, reasoning (main) | 0.86  (132/154) | 0.88  (120/136) | 0.67  (12/18) |

Discordant candidates are those on which the CEM verdict and the first-pass judgment differ; values are accuracy against the LLM judge (correct/total). The Adjudicator accepted 140 of the 154 candidates with reasoning (133 and 7 in the two groups) and 113 without reasoning (112 and 1). Candidate-level records are in S17.

**S16. Agreement of final NCO sets with the main configuration**

| **Configuration** | **Empagliflozin vs. sitagliptin** | **Warfarin vs. apixaban** | **Lisinopril vs. HCTZ** | **Pooled** |
| --- | --- | --- | --- | --- |
| Adjudicator, no reasoning | 0.97 (0.97) | 0.90 (0.92) | 0.79 (0.87) | 0.89 (0.92) |
| Reasoning in all agents | 0.84 (0.87) | 0.83 (0.85) | 0.81 (0.87) | 0.83 (0.87) |
| Curator: ingredient names only | 0.85 (0.88) | 0.89 (0.91) | 0.86 (0.91) | 0.87 (0.90) |
| Curator: raw label text | 0.94 (0.96) | 0.87 (0.90) | 0.83 (0.88) | 0.88 (0.91) |
| No concept hierarchy | 0.90 (0.92) | 0.93 (0.95) | 0.87 (0.91) | 0.90 (0.93) |

Jaccard index of the final NCO sets (proportion of identical decisions in parentheses). HCTZ, hydrochlorothiazide; NCO, negative control outcome.

**S17. All discordant candidates in the main configuration** — Supplementary_Data.xlsx, sheet S17.

**S18. Decision paths and misclassified candidates in the main configuration**

**(a) Concordance between CEM and the first-pass judgment, location of false positives, and adjudication load**

|  | **Empagliflozin vs. sitagliptin** | **Warfarin vs. apixaban** | **Lisinopril vs. HCTZ** | **Pooled** |
| --- | --- | --- | --- | --- |
| Candidates | 117 | 115 | 117 | 349 |
| Concordant, n (%) | 91 (77.8%) | 47 (40.9%) | 57 (48.7%) | 195 (55.9%) |
| Both accept / FP | 66 / 15 | 23 / 0 | 14 / 2 | 103 / 17 |
| Both reject / FN | 25 / 2 | 24 / 6 | 43 / 13 | 92 / 21 |
| Discordant (routed to adjudication), n (%) | 26 (22.2%) | 68 (59.1%) | 60 (51.3%) | 154 (44.1%) |
| CEM rejects, first pass accepts | 18 | 61 | 57 | 136 |
| CEM accepts, first pass rejects | 8 | 7 | 3 | 18 |
| Accepted by Adjudicator / FP | 20 / 5 | 64 / 4 | 56 / 9 | 140 / 18 |
| False positives: concordant / discordant | 15 / 5 | 0 / 4 | 2 / 9 | 17 / 18 |

FP and FN are relative to the LLM judge. CEM, Common Evidence Model; FN, false negative; FP, false positive; HCTZ, hydrochlorothiazide.

**(b) Classification of misclassified candidates by decision path**

| **Code** | **Concordant** | **Adjudicated** | **Total** |
| --- | --- | --- | --- |
| False positives, n | 17 | 18 | 35 |
| Error type |  |  |  |
| Type 1 (label-grounded, direct) | 0 | 0 | 0 |
| Type 2 (label-grounded, indirect) | 12 | 12 | 24 |
| one inference step | 9 | 2 | 11 |
| two or more steps | 3 | 10 | 13 |
| Type 3 (off-label, established) | 0 | 5 | 5 |
| Type 4 (speculative) | 5 | 1 | 6 |
| Failure mode |  |  |  |
| Label dismissed as non-causal | 3 | 1 | 4 |
| Label override | 0 | 2 | 2 |
| No downstream reasoning | 3 | 3 | 6 |
| Taxonomic mismatch | 4 | 0 | 4 |
| Mechanism-first filtering | 2 | 7 | 9 |
| Knowledge gap | 3 | 4 | 7 |
| Other | 2 | 1 | 3 |
| Broader term present in hierarchy input | 7 | 1 | 8 |
| False negatives, n | 21 | 4 | 25 |
| Over-extended mechanism | 15 | 3 | 18 |
| Label misreading | 5 | 0 | 5 |
| Indication confusion | 1 | 0 | 1 |
| CEM evidence over-weighted | 0 | 0 | 0 |
| Other | 0 | 1 | 1 |

Errors are relative to the LLM judge. Concordant: CEM and the first-pass judgment agreed (deciding rationale: Suggester); adjudicated: decided by the Adjudicator. Codes were assigned by an LLM with a fixed codebook. Definitions are given in Section 3.2.7(d) and Supplementary S9.

**S19. Efficiency by agent and configuration (three drug pairs combined)**

| **Configuration** | **Agent** | **Calls** | **Input tokens** | **Output tokens** | **Reasoning tokens** | **LLM time (min)** |
| --- | --- | --- | --- | --- | --- | --- |
| Main | Curator | 6 | 23,971 | 2,388 | 0 | 1.1 |
|  | Suggester | 349 | 576,517 | 46,488 | 0 | 21.4 |
|  | Adjudicator | 154 | 384,264 | 326,874 | 307,725 | 129.8 |
|  | Diagnostician | 3 | 6,175 | 444 | 0 | 0.2 |
|  | Wall-clock total (CEM waiting) |  |  |  |  | 158 (2.6) |
| Adjudicator, no reasoning | Curator | 6 | 23,971 | 2,388 | 0 | 1.1 |
|  | Suggester | 349 | 576,517 | 46,488 | 0 | 21.4 |
|  | Adjudicator | 154 | 384,572 | 23,441 | 0 | 11.3 |
|  | Diagnostician | 3 | 5,994 | 418 | 0 | 0.2 |
|  | Wall-clock total (CEM waiting) |  |  |  |  | 36 (1.9) |
| Reasoning in all agents | Curator | 6 | 23,959 | 31,213 | 28,503 | 12.4 |
|  | Suggester | 349 | 619,629 | 620,395 | 576,900 | 245.0 |
|  | Adjudicator | 177 | 464,676 | 367,752 | 345,457 | 146.2 |
|  | Diagnostician | 3 | 6,653 | 9,862 | 9,460 | 3.9 |
|  | Wall-clock total (CEM waiting) |  |  |  |  | 410 (1.9) |
| Curator: ingredient names only | Suggester | 351 | 313,691 | 49,698 | 0 | 21.1 |
|  | Adjudicator | 158 | 268,046 | 347,700 | 328,042 | 136.9 |
|  | Diagnostician | 3 | 3,808 | 437 | 0 | 0.2 |
|  | Wall-clock total (CEM waiting) |  |  |  |  | 161 (3.0) |
| Curator: raw label text | Suggester | 349 | 2,892,288 | 48,216 | 0 | 35.7 |
|  | Adjudicator | 155 | 1,366,845 | 281,117 | 261,992 | 119.3 |
|  | Diagnostician | 3 | 26,159 | 453 | 0 | 0.3 |
|  | Wall-clock total (CEM waiting) |  |  |  |  | 160 (3.1) |
| No concept hierarchy | Curator | 6 | 23,971 | 2,388 | 0 | 1.1 |
|  | Suggester | 349 | 468,256 | 45,559 | 0 | 20.4 |
|  | Adjudicator | 163 | 352,888 | 363,240 | 342,926 | 143.7 |
|  | Diagnostician | 3 | 6,194 | 426 | 0 | 0.2 |
|  | Wall-clock total (CEM waiting) |  |  |  |  | 169 (2.8) |

The Curator makes no LLM calls in the names-only and raw-label configurations. Calls include retries. Output tokens include reasoning tokens. LLM time is the sum of the response times of the LLM calls; the last row of each configuration gives the wall-clock time of the whole run, with the time spent waiting for the CEM WebAPI in parentheses. CEM, Common Evidence Model; LLM, large language model.

**S20. Cost–accuracy trade-off across configurations**

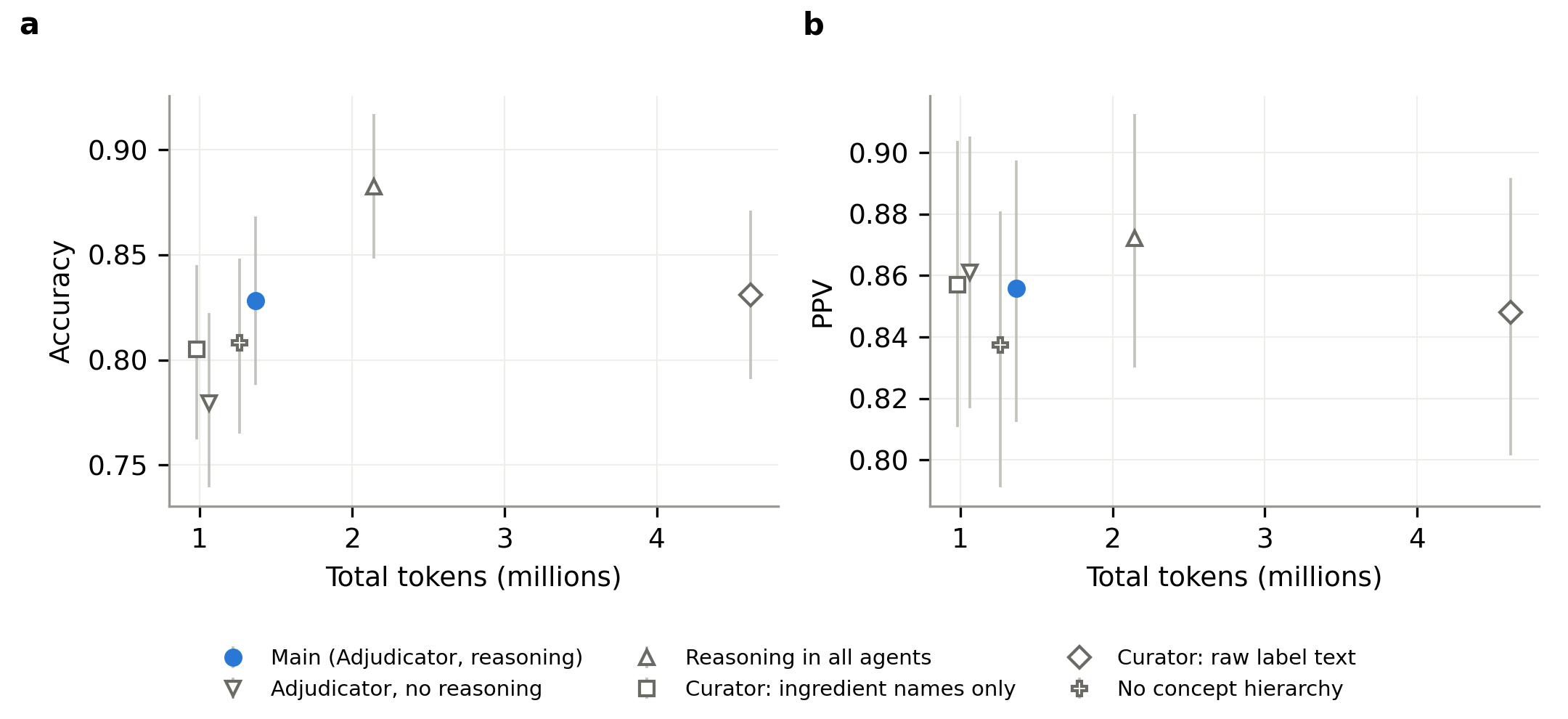

Total tokens (three drug pairs combined) against pooled (a) accuracy and (b) PPV for each MENTOR configuration. Error bars are bootstrap 95% CIs. CI, confidence interval; PPV, positive predictive value.

**S21. Availability and threshold analysis**

**(a) Availability of CEM-proposed outcomes in AUSOM**

| **Drug pair** | **CEM proposed** | **In AUSOM** | **≥1,000** | **≥5,000** | **≥10,000** |
| --- | --- | --- | --- | --- | --- |
| Empagliflozin vs. sitagliptin | 227 | 149 (65.6%) | 69 (30.4%) | 33 (14.5%) | 20 (8.8%) |
| Warfarin vs. apixaban | 169 | 106 (62.7%) | 35 (20.7%) | 20 (11.8%) | 14 (8.3%) |
| Lisinopril vs. HCTZ | 170 | 100 (58.8%) | 29 (17.1%) | 16 (9.4%) | 12 (7.1%) |

Final ATLAS proposal list (after its optimization step); patient counts from the full AUSOM CDM, counted with descendants, without the exclusion list. AUSOM, Ajou University School of Medicine; CDM, common data model; CEM, Common Evidence Model; HCTZ, hydrochlorothiazide.

**(b) Number of NCOs and MENTOR PPV by patient-count threshold (main configuration)**

| **Patient-count threshold** | **Drug pair** | **Candidates** | **MENTOR NCOs** | **CEM NCOs** | **Judge-valid** | **MENTOR PPV** |
| --- | --- | --- | --- | --- | --- | --- |
| 70,000 | Empagliflozin vs. sitagliptin | 6 | 1 | 2 | 0 | 0.00 |
|  | Warfarin vs. apixaban | 6 | 5 | 1 | 6 | 1.00 |
|  | Lisinopril vs. HCTZ | 6 | 2 | 0 | 1 | 0.50 |
| 50,000 | Empagliflozin vs. sitagliptin | 11 | 6 | 6 | 5 | 0.83 |
|  | Warfarin vs. apixaban | 11 | 9 | 1 | 11 | 1.00 |
|  | Lisinopril vs. HCTZ | 11 | 4 | 0 | 6 | 0.75 |
| 30,000 | Empagliflozin vs. sitagliptin | 20 | 11 | 12 | 9 | 0.73 |
|  | Warfarin vs. apixaban | 20 | 15 | 3 | 18 | 1.00 |
|  | Lisinopril vs. HCTZ | 20 | 9 | 0 | 12 | 0.89 |
| 20,000 | Empagliflozin vs. sitagliptin | 49 | 35 | 34 | 26 | 0.69 |
|  | Warfarin vs. apixaban | 47 | 37 | 15 | 40 | 1.00 |
|  | Lisinopril vs. HCTZ | 48 | 25 | 6 | 28 | 0.88 |
| 15,000 | Empagliflozin vs. sitagliptin | 75 | 55 | 50 | 44 | 0.76 |
|  | Warfarin vs. apixaban | 73 | 59 | 22 | 62 | 0.98 |
|  | Lisinopril vs. HCTZ | 75 | 46 | 8 | 47 | 0.87 |
| 10,000 | Empagliflozin vs. sitagliptin | 117 | 86 | 74 | 69 | 0.77 |
|  | Warfarin vs. apixaban | 115 | 87 | 30 | 90 | 0.95 |
|  | Lisinopril vs. HCTZ | 117 | 70 | 17 | 74 | 0.84 |

Reconstructed from the main runs (candidates with a CEM record whose patient count met the threshold). NCO counts are raw counts, before parent–child or near-synonymous concepts are counted as one. CEM, Common Evidence Model; HCTZ, hydrochlorothiazide; NCO, negative control outcome; PPV, positive predictive value.

**S22. Representative errors by failure mode**

| **Failure mode** | **Drug pair** | **Outcome (path)** | **What the deciding agent missed** |
| --- | --- | --- | --- |
| Label dismissed as non-causal | Empagliflozin vs. sitagliptin | Acute pharyngitis (concordant, FP) | Upper respiratory tract infection and nasopharyngitis are listed; the Suggester named them but attributed them to chance and accepted the outcome. |
| Taxonomic mismatch | Empagliflozin vs. sitagliptin | Acute sinusitis (concordant, FP) | Upper respiratory tract infection is listed and was in the hierarchy input; the Suggester did not link sinusitis to it and called the outcome unlabeled. |
| Mechanism-first filtering | Lisinopril vs. HCTZ | Closed fracture of rib (adjudicated, FP) | Hypotension, orthostatic hypotension, and dizziness are listed; the Adjudicator reasoned from the drugs' mechanisms without considering falls (two inference steps). |
| No downstream reasoning | Empagliflozin vs. sitagliptin | Disorders of initiating and maintaining sleep (concordant, FP) | Nocturia is listed for empagliflozin; the Suggester named it but rejected the one-step link to disturbed sleep maintenance. |
| Label override | Lisinopril vs. HCTZ | Acute bronchitis (adjudicated, FP) | Cough is listed for lisinopril; the Adjudicator noted that the parent-level CEM label evidence likely reflected cough but accepted the outcome without exact-level evidence. |
| Knowledge gap | Warfarin vs. apixaban | Osteoarthritis of knee (adjudicated, FP) | Not in the context; the judge cites inhibition of vitamin K-dependent matrix Gla protein and observational data comparing vitamin K antagonists with direct oral anticoagulants. |
| Over-extended mechanism | Warfarin vs. apixaban | Chronic gastritis (concordant, FN) | Rejected because anticoagulant-related bleeding 'can manifest as or exacerbate gastritis', an indirect pathway; the judge accepted the outcome. |

HCTZ, hydrochlorothiazide; FP, false positive; FN, false negative.
